# Prevalence and factors associated with low utilization of maternal services among women attending health services in Marávia and Zumbo districts, Tete province, Mozambique

**DOI:** 10.1101/2025.03.04.25323363

**Authors:** Maria Suzana Maguele, Emídio Cumbane, Jorda Daneluzzo, Málica de Melo, Vânia Benzane, Fídel Paizone, Benilde Homo, Anna Galle

## Abstract

Worldwide, Maternal mortality continues to be a major problem. This study explored the prevalence and factors associated with low utilization of maternal services among women attending health services in Marávia and Zumbo districts, Tete province, Mozambique. Using a probability proportional sampling, data of 381 participants were collected using an interview-administered questionnaire. Binary and multivariate logistic regression analyses were performed. Of the 381 participants 283 (74,3%), (95% CI: 69.7–78.5) had four or more ANC visits, and 98 (25,7%) had less. More than a half of participants 211 (56,4%) initiated ANC visit after the first trimester of pregnancy, and 163 (43,6%) (95% CI: 38.6–48.6) within the first trimester, 267 (70.1%) (95% CI: 65.4–74.5) had delivered at health facility and 117(29.9%) delivered outside. 275 (72,2%) (95% CI: 65.4–74.5) had three or more PNC, and 106 (27,8%), had less. There was association between the less than four ANC and the age of women (AOR, 0. 007, 95% CI: 0.142−0.735, p = 0.007), delaying more than hour to reach the health facility (AOR, 2. 517, 95% CI: 1.327−4.773, p = 0.005), and weak knowledge on the frequency of ANC (AOR, 2. 455, 95% CI: 1.47−4.099, p = 0.001). There was association between not attending institutional delivery and the women’s lack of commitment with religion (AOR, 3. 056, 95% CI: 1.332−7.012, p = 0.008), women’s primary education level (AOR, 5. 635, 95% CI: 1.114−28.505, p = 0.037) and women not satisfied with ANC services (AOR, 2. 257, 95% CI: 1.174−4.341, p = 0.015). There was association between less than three PNC visits and women status of married (AOR, 0. 040, 95% CI: 0.015−0.104, p = 0.001). There is an urgent need for strategies to improve access to health services, by removing socio economic, cultural and structural barriers.

## Introduction

Worldwide, Maternal mortality continues to be a major problem, with a woman dying from complications related to pregnancy and birth.

Global estimates indicated a maternal mortality rate (MMR), of 223 maternal deaths per 100 000 live births, in 2020. In terms of ranking, Sub-Saharan Africa accounts for the highest MMR estimated at 545 maternal deaths per 100 000 live births making up for a substantial proportion in these global estimates, of 87_90% [1]

Mozambique is a one of the Sub-Saharan countries with an extremely high national MMR, although reductions estimated at 452 and 233 maternal deaths per 100,000 live births have been recorded, from 2017 to 2023, respectively [2–4].

While Mozambique is committed with the sustainable development goals (SDG) to reduce the current estimates, the challenge over the next seven years is to work towards reducing the maternal mortality rate by no more than 140/100 000 live births, by 2030 [5].

Evidence has shown that utilization of appropriate maternal health services, indicated as regular antenatal care (ANC) attendance, professionally skilled births or institutional delivery (ID) and post-natal care (PNC) visits, are contributing factors to improve maternal outcomes and prevent deaths[5–7]. For instance, the use of ANC, institutional delivery and PNC services have proven to reduce maternal and child mortality and morbidity by the means of early preventative measures, including health promotion, counselling on the use of institutional delivery, planning and readiness for birth and care after delivery [8–14].

With good antenatal care, some of the risk factors affecting pregnant woman, for example, HIV (Human Immunodeficiency Virus), Malaria, hypertension and malnutrition can be detected and treated timeously [15, 16]. Prompt treatment of such condition can be a contributing factor for preventing maternal and child morbidity and low birth weight in countries where HIV, malaria, malnutrition are still a high burden [17–20].

World Health Organization (WHO), recommends optimising opportunities to screening, prevention and treatment of conditions during pregnancy, through maximizing contact and interactions between health providers and pregnant women [21]. Monthly visits until the end of pregnancy is expected for every woman, however, local context can vary, and adaptation may need to be implemented [22].

Based on the WHO recommendations on the local contextual ANC basic and focused package, the Mozambican Ministry of health recommends for reduction of maternal complications a minimum of four antenatal visits during pregnancy, and the first visit being within the first three months [22–24].

The Delivery is recommended to be with a skilled health provider at a health facility. Women should be allowed to have at least four PNC visits within the 42 days after the delivery. A first PNC being within 48 hours after delivery [22, 24–26]. In Mozambique, the first visit after childbirth is done immediately before the leave of maternity [23].

Although there is evidence regarding utilisation of maternal services with increased maternal outcomes, utilization of maternal services is still low in most settings, especially in sub-Saharan Africa, where practices of home delivery, with no assistance from a skilled individual are strong and rooted [27–30]. Prevalence of utilization of ANC, ID and PNC varies across countries where data is available, with numbers accounting for around 55.2%, 78.8%, and 40%, respectively[30]. For example, evidence from Ethiopia indicates a high prevalence (70%) of home deliveries[16, 30]. Another study from somewhere indicated only 19% of women attended at least four ANC while 7% attended PNC [8].

According to the Mozambican National Health demographic Survey (NHDS), (2023), only 49% of women who had children at least two years before the survey, had four or more ANC visits during their pregnancy. Around 65% of women had births with a skilled provider at a health facility but only 36 % had PNC visits within the first 2 days after delivery [4]. Another study undertaken in a rural area, indicated only 11% of ANC visits in the first three months of pregnancy [31].

Among factors hindering women’s utilization of maternal services, there are factors related to their low socio-economic status, such as long walk distances from health facilities, lack of transportation to health facility, lack knowledge and preparation on pregnancy emergencies as well as a low educational level [27, 32, 33].

In Mozambique as in many others African countries, the lack of employment resulting in poverty impact women’s ability in accessing health services during pregnancy [33]. Socio-cultural and structural determinants such as marital status of women, literacy, availability, as well as the perspective and satisfaction with the health services limit the use of maternal services [27, 32–36]. Generally in Mozambique, women from urban areas (where individuals may have better access to resources and infrastructure) are more likely to attend ANC services, have ID and attend more PNC services compared to those from rural areas, where low education and the incidence of no formal work tends to be the reality [4]. Determinants of maternal health services use are likely to be a combination of individual, socio-cultural and structural factors. The low educational and economic levels as well as the social gender norms favourable to male dominance may be the key contributing factors for the low use of health services during pregnancy, birth, and postpartum [4, 6, 11, 27, 32–34, 36].

This study focused on women who have used maternal services in two districts of Tete province, Mozambique.

Mozambique is an African country committed in the efforts to archive the United Nations (UN) sustainable goals; however, the country is still grappling with a lack of evidence reporting the prevalence of maternal service utilization focusing on the three key indicators of maternal services, namely ANC, ID and PNC. Available Data is mostly reported from National Health demographics data based on households’ surveys, which analysed other variables reported on descriptive statistics. Other available data is from studies which measured only the use of ANC services [4, 31]. Hence the variables, methods and measures may be different. There still a gap in information regarding factors associated with low utilization of the three maternal services among those who have utilized it.

More up to date Information is urgently required to suggest specific interventions that address the needs of women in utilizing maternal services in specific settings. This study will provide evidence required to encourage policymakers to act towards achieving SDG 3 and 5[23].

With this study we aimed to determine the prevalence and factors associated with low utilization of maternal services (ANC, ID, PNC), among women attending maternal services at health facilities in the Marávia and Zumbo districts, Tete province, Mozambique.

## Material and methods

### Study area and setting

The study was conducted in six health facilities in Marávia and Zumbo Districts, Tete. The selected health facilities comprised one District level health facility and two rural health facilities in each District. Tete province is located in the northern area of Mozambique, covers an area of 98,417 km² and has a population of approximately 2,648,941 inhabitants, where about 13.6% live in urban areas [15]. The number of births in this setting was 2,906 in 2019 and the province held the fifth position for higher national rates of maternal death and other maternal indicators [2, 4, 23]. The abovementioned Districts were selected to implement a program to improve maternal health. The research was conducted in the perspective of an institutional baseline to understand the prevalence of maternal service utilization and its contributing factors. We determined the prevalence of low utilization of maternal services and accessed factors associated.

### Study design and population

This was a cross-sectional study that explored prevalence of low utilization of maternal services and the factors associated among maternal service users in Tete.

#### Study Population

The study population were women of reproductive age, who had given last birth within 1 year at moment of data collection and attending post-natal services in one of the six selected health facilities in October and November 2023. In Mozambique, although the high rates of unemployment majorly affect women, in rural areas, women with children are likely to be away from home during day, for farm or other informal work/activities to provide subsistence for their children [37].

#### Study sample

The sample size calculation was an institutional-based size sample from postnatal service users, using a 95% confidence interval and a 5% degree of precision. We therefore added 10% to the sample for possible invalid responses. The systematic sampling strategy (k=13) was employed to select the 381 participants from all the health facilities, from the 23^rd^ of October to the 17^th^ of November 2023. The systematic selection provided an equal chance for all the potential participants to be selected so that the findings could be generalized to similar populations in the services.

To be selected participants should have been in line with the following criteria: Had given last birth within 1 year at moment of data collection.

Attending post-natal services in one of the six selected health facilities during the period of data collection.

Had a live birth (last delivery)

Autonomy to consent to participation or the ability to obtain consent from their guardians (A minor under 18 years of age)

### Data collection

#### Instruments

The questionnaire for the survey was developed based on the theoretical model for utilization of maternal services (Anderson, 1973) [38, 39], covering sociodemographic; obstetric history, knowledge, satisfaction, utilization of services during pregnancy, delivery and post-natal.

The selection of the questions was designed to address the socio-cultural context of women who have ever had children regarding their utilization of maternal services during their pregnancy, delivery as well as post-natal care.

The questionnaire was designed to enable the estimation of the prevalence of utilization of maternal services and the factors associated with low utilization of maternal services.

The Mozambican ministry of health, following the WHO recommendations, has adapted recommendations for maternal service utilisation, being, a monthly visit during antenatal period with a minimum of four antenatal visits and the first visit being early within the first trimester (twelve weeks of pregnancy); a skilled/professional institutional delivery and a minimum of four post-natal visits within 42 days after delivery, first within the first 48 hours, a second within the first 7 days, a third within 28 days and fourth within 42 days. For the best understanding, in this study, we adapted our measure to three PNC visits after the leave from maternity. The latter is due to the first PNC visit occurring during internal care after delivery and before leaving the maternity. This would have been difficult to measure since the information was self-reported. We assumed every woman who had any contact with maternal services after birth, had at least one PNC visit.

#### Measures

Utilization of maternal services was measured as follows:

ANC services utilization-number of visits across pregnancy and time of first visit. Comparison was made between those who attended less or more than four ANC visits; and comparison between those who had their first ANC visit after or within the first three months of pregnancy.

ID services utilization-number of non-institutional vs institutional deliveries. Comparison was made between those who used health facilities for delivery regardless of location and type of health facility and those not using ID.

PNC services utilization-number of visits within the 42 days following delivery. Comparison was made between those who attended less than three and those attended three or more PNC visits.

Dependant variables comprised utilization of maternal services (ANC, ID and PNC) and were obtained by asking participants the following questions:

- During your last pregnancy, how many times did you visit the ANC services with the purpose of taking care of your pregnancy?
- How far were you with the pregnancy, (in months) when you first made use of ANC services for care?
- Where did you have your delivery (in this hospital, any other hospital, home, neighbourhood, on your way to the hospital)?
- How many times did you return to PNC services to check on you and your newborns health in the following 42 days after you gave birth?

Independent variables-Were informed by the available literature and theoretical model to explain the factors associated with utilization of maternal health. In addition, the contextual socio-cultural factors in Mozambique which may explain the low utilization of maternal health were also included. The independent variables comprised two sections: one on socio-demographic characteristics and obstetric history measured as categorical variables and section two investigated the knowledge of and importance given to maternal services. To measure the knowledge, participants responses were categorized into two categories. Those with knowledge were those indicated by any correct response and those with no knowledge, were those indicated any incorrect response. The Socio-demographic factors included: Age; Employment status of the respondent; education level; Relationship status of respondent; Commitment to religion; number of household members; Educational level of the partner; Employment status of the partner; number of living children; household monthly financial income; distance between residence and the nearest hospital; time to reach the nearest hospital; means for transport to hospital; place where they get information on sexual and reproductive health; who makes the decision on sexual and reproductive health including the use of family planning; knowledge and importance about maternal health issues and satisfaction with services during the last use of maternal services. The questions presented to participants to assess the prevalence and factors associated with low utilization of maternal services are detailed in the questionnaire S1 File.

### Pilot

Nearly 10% of sample, was piloted in a health facility with a similar setting to ensure clarity of the questions and consistency in the methods of questioning and the data collection procedure. After the pilot, few questions relating to the utilization of health services and obstetric history were re-formulated to be more adequate for the women attending rural settings in the Districts of Marávia and Zumbo.

### Ethics

This study was approved by the Interinstitutional Health Bioethics Committee of Tete (CIBS-Tete), reference: /CIBSTete/2023. Administrative Permission was obtained from the Mozambican Ministry of Health and local government at provincial and District levels. Participants were recruited after their attendance in post-natal care services. Consent forms and assent forms were explained and distributed to all participants before the study. All participants provided written informed consent voluntarily. Participants under 18 years of age provided assent and the consent from their parents or guardians. No monetary reimbursement was given to them for their participation. Anonymity and confidentiality were ensured, and the participants’ names were not written on any questionnaires. Privacy was maintained by keeping the participants separated from the local of attendance during the completion of the interview-administered questionnaire. They were told that their participation was voluntary and that they had the right to terminate it, and they were assured that they would not be affected in any way if they decided to do this. There was minimal risk that the study had the potential to bring back negative memories regarding their negative or stressful experiences of pregnancy and delivery, but participants were told to inform the researchers if this occurred, and they were assured of the availability of referral services for psychological assistance. Participants benefited from the information about maternal health, how to take care of themselves during pregnancy, delivery, and post-natal period and how to prevent maternal and child morbidity. The researcher made available contacts for reference services for assistance in case participants needed help if they experience stress or negative feelings resulting from participating in the study. No participants reported such needs.

### Data analysis

After assessing data completeness, the data was analysed using SPSS version 20.0 statistical software. Proportions were used to estimate:

- Prevalence of less than four ANC visits during pregnancy.
- Prevalence of first ANC visit after the first trimester of pregnancy.
- Prevalence of non-ID.
- Prevalence of less than three PNC visits within 42 days following delivery.

Logistic regression was used to identify less than four ANC visits, factors associated with first ANC visit after first trimester, factors associated with non-institutional delivery, and factors associated with less than three PNC visits. Odds ratios (OR) and 95% confidence intervals (CI) are reported. After conducting the simple logistic regression of low utilization of services and the potential risk factors, significant risk factors, were then included in the multivariable logistic regression. For this study, a p value *<*0.05 was deemed statistically significant.

## Results

### Sample description

We had an initial estimated sample size of 356 participants. During the data collection period, 394 participants were enrolled. After data cleanness, 13 questionnaires (corresponding to 3%) were removed due to incomplete responses. A total of 381(an increase of 7%) questionnaires were included for analysis and results are presented. Table 1 presents the sociodemographic characteristics of participants.

**Table 1.**
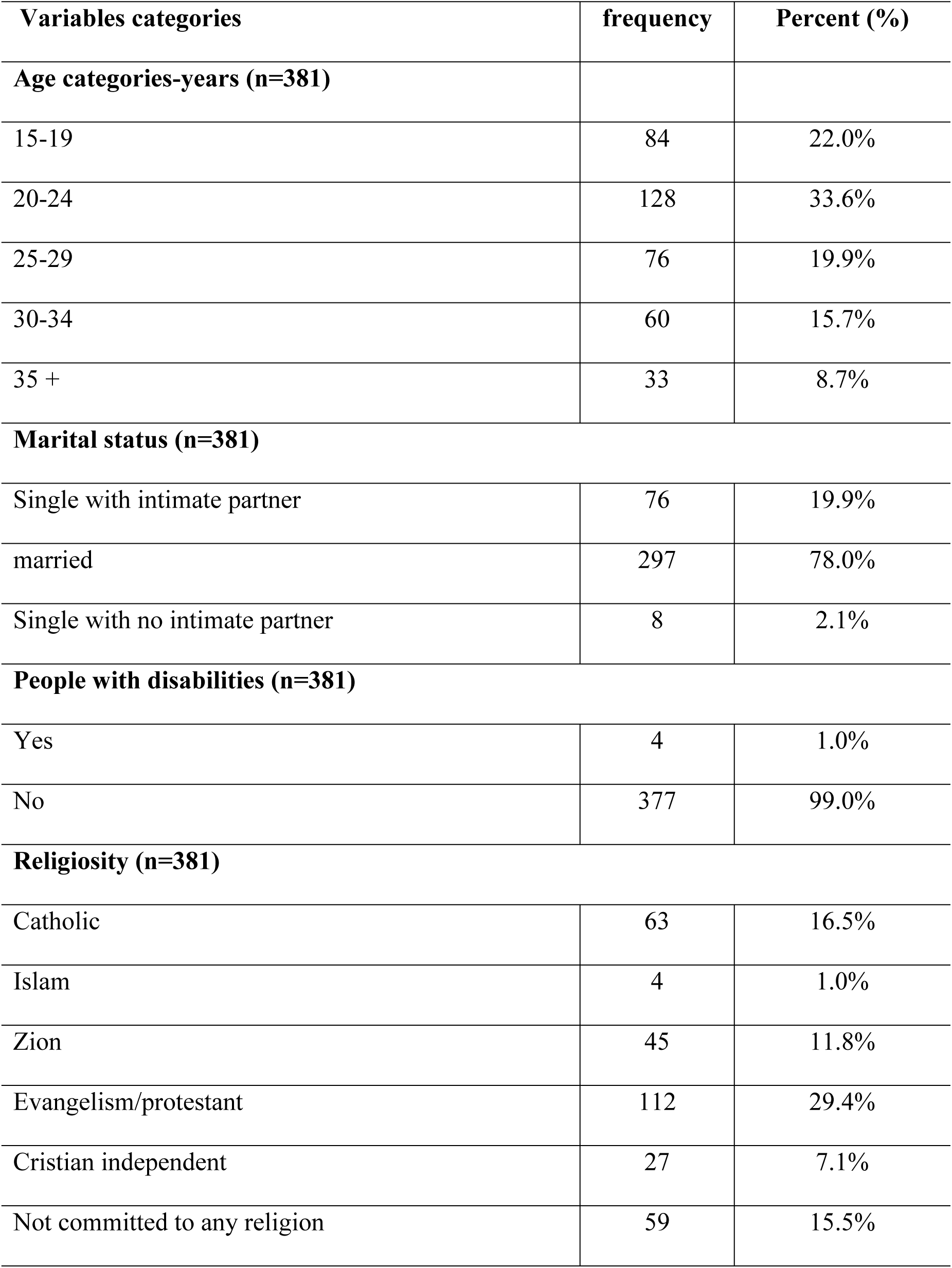
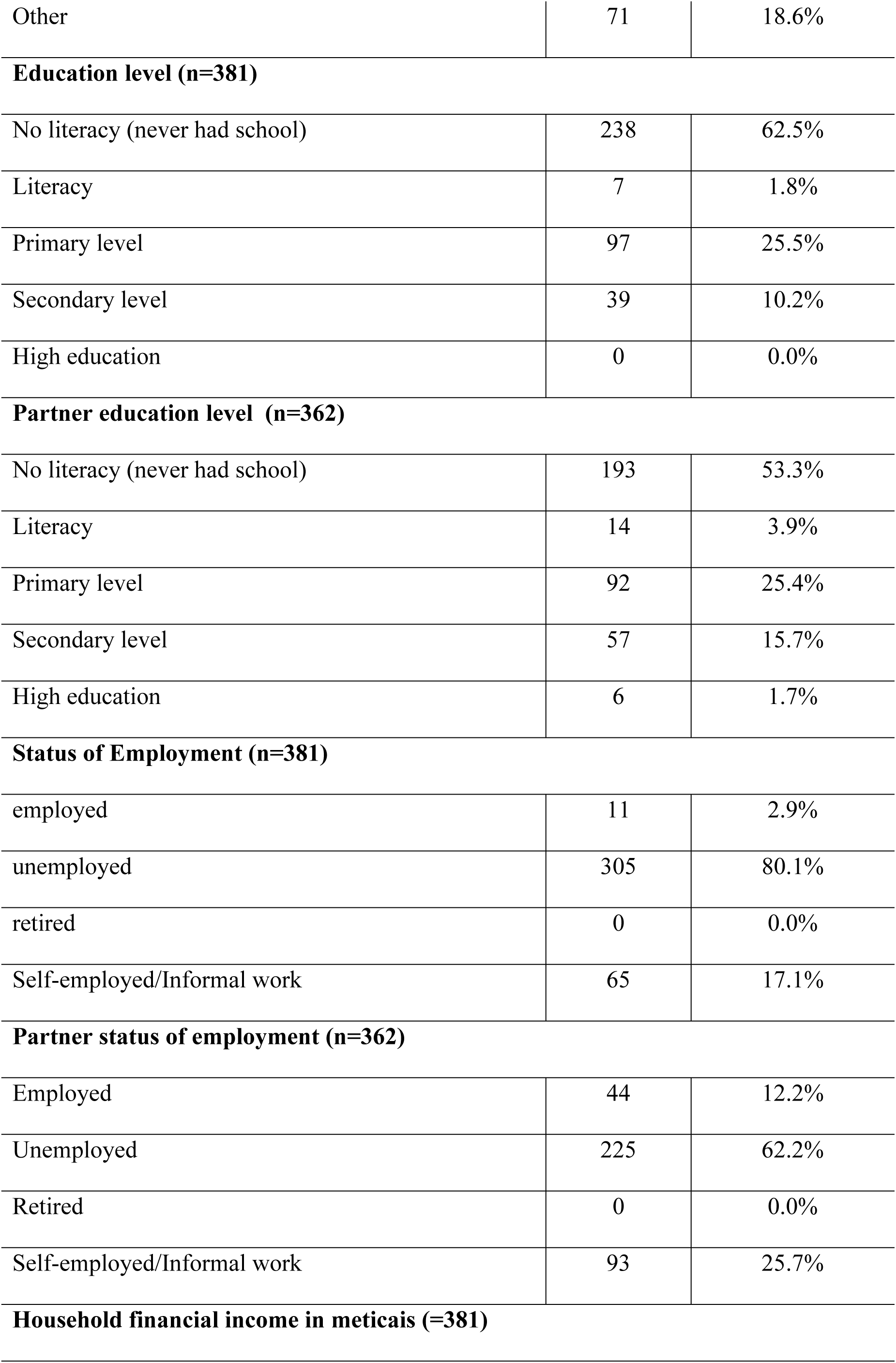
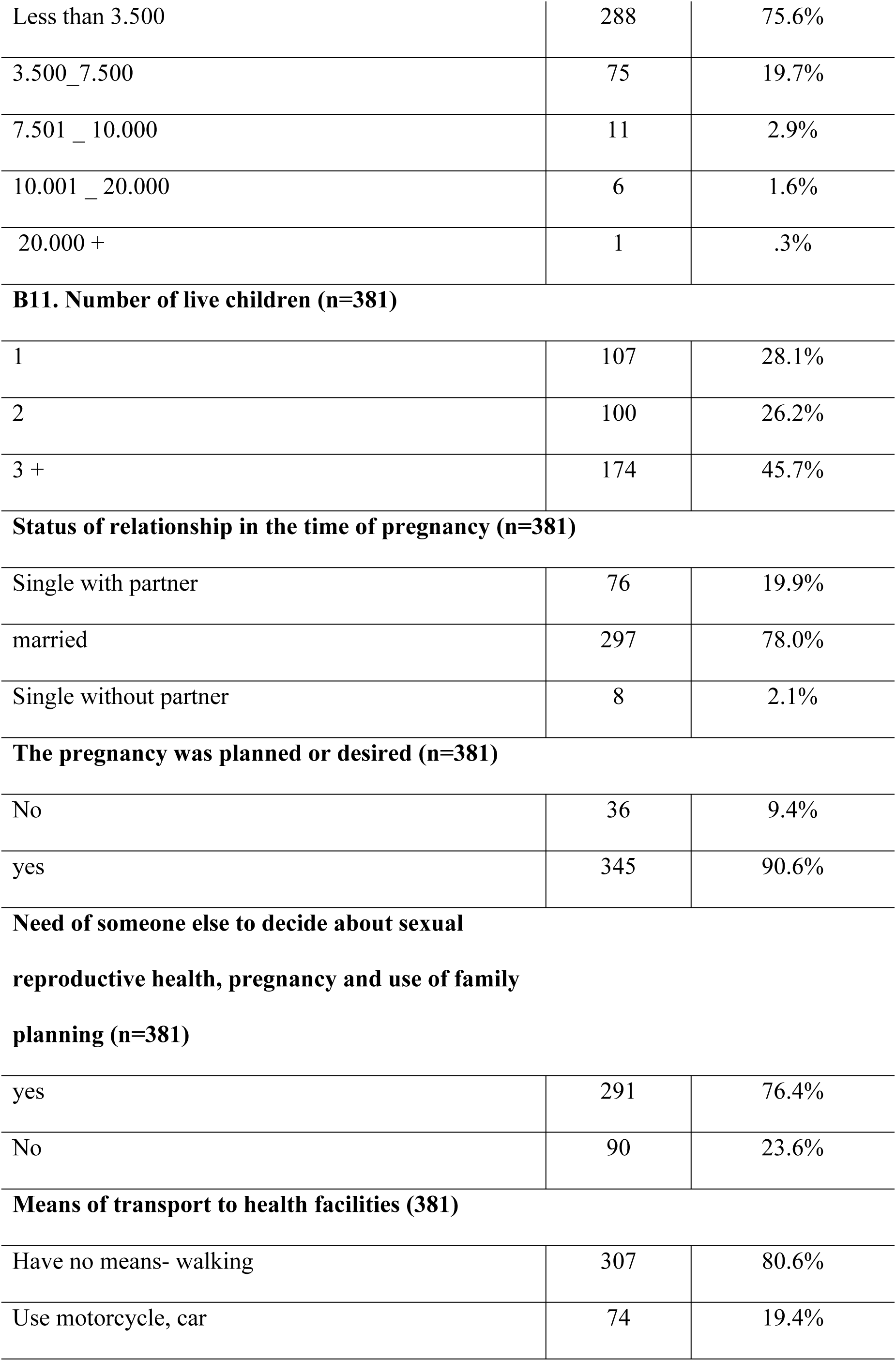
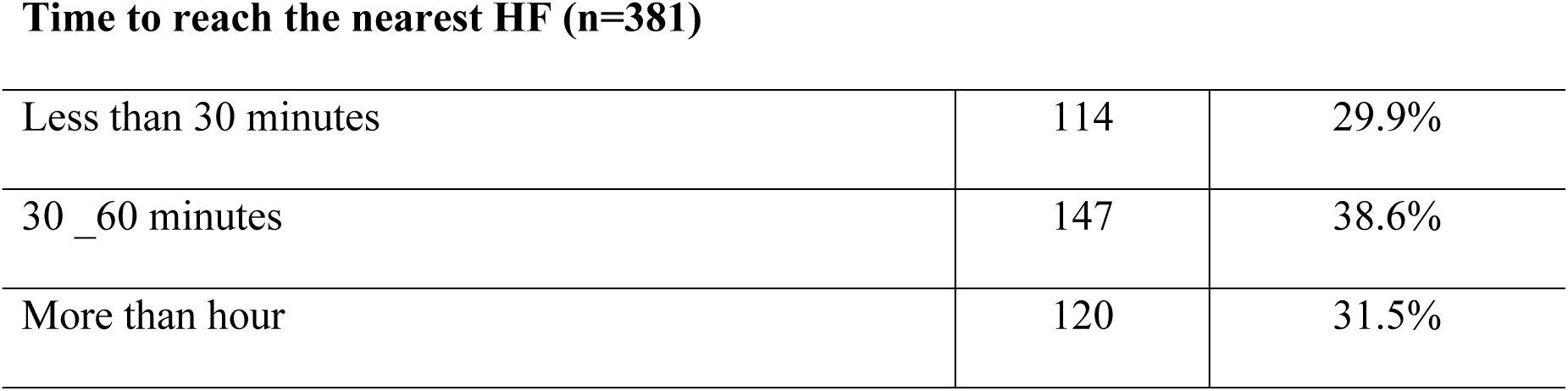
Sociodemographic characteristics of participants.

| **Variables categories** | **frequency** | **Percent (%)** |
| --- | --- | --- |
| **Age categories-years (n=381)** |  |  |
| 15-19 | 84 | 22.0% |
| 20-24 | 128 | 33.6% |
| 25-29 | 76 | 19.9% |
| 30-34 | 60 | 15.7% |
| 35 + | 33 | 8.7% |
| **Marital status (n=381)** |  |  |
| Single with intimate partner | 76 | 19.9% |
| married | 297 | 78.0% |
| Single with no intimate partner | 8 | 2.1% |
| **People with disabilities (n=381)** |  |  |
| Yes | 4 | 1.0% |
| No | 377 | 99.0% |
| **Religiosity (n=381)** |  |  |
| Catholic | 63 | 16.5% |
| Islam | 4 | 1.0% |
| Zion | 45 | 11.8% |
| Evangelism/protestant | 112 | 29.4% |
| Cristian independent | 27 | 7.1% |
| Not committed to any religion | 59 | 15.5% |
| Other | 71 | 18.6% |
| **Education level (n=381)** |  |  |
| No literacy (never had school) | 238 | 62.5% |
| Literacy | 7 | 1.8% |
| Primary level | 97 | 25.5% |
| Secondary level | 39 | 10.2% |
| High education | 0 | 0.0% |
| **Partner education level (n=362)** |  |  |
| No literacy (never had school) | 193 | 53.3% |
| Literacy | 14 | 3.9% |
| Primary level | 92 | 25.4% |
| Secondary level | 57 | 15.7% |
| High education | 6 | 1.7% |
| **Status of Employment (n=381)** |  |  |
| employed | 11 | 2.9% |
| unemployed | 305 | 80.1% |
| retired | 0 | 0.0% |
| Self-employed/Informal work | 65 | 17.1% |
| **Partner status of employment (n=362)** |  |  |
| Employed | 44 | 12.2% |
| Unemployed | 225 | 62.2% |
| Retired | 0 | 0.0% |
| Self-employed/Informal work | 93 | 25.7% |
| **Household financial income in meticaís (=381)** |  |  |
| Less than 3.500 | 288 | 75.6% |
| 3.500_7.500 | 75 | 19.7% |
| 7.501 _ 10.000 | 11 | 2.9% |
| 10.001 _ 20.000 | 6 | 1.6% |
| 20.000 + | 1 | .3% |
| **B11. Number of live children (n=381)** |  |  |
| 1 | 107 | 28.1% |
| 2 | 100 | 26.2% |
| 3 + | 174 | 45.7% |
| **Status of relationship in the time of pregnancy (n=381)** |  |  |
| Single with partner | 76 | 19.9% |
| married | 297 | 78.0% |
| Single without partner | 8 | 2.1% |
| **The pregnancy was planned or desired (n=381)** |  |  |
| No | 36 | 9.4% |
| yes | 345 | 90.6% |
| **Need of someone else to decide about sexual reproductive health, pregnancy and use of family planning (n=381)** |  |  |
| yes | 291 | 76.4% |
| No | 90 | 23.6% |
| **Means of transport to health facilities (381)** |  |  |
| Have no means- walking | 307 | 80.6% |
| Use motorcycle, car | 74 | 19.4% |
| Time to reach the nearest HF (n=381) |  |  |
| Less than 30 minutes | 114 | 29.9% |
| 30 _60 minutes | 147 | 38.6% |
| More than hour | 120 | 31.5% |

The age of participants varied from 15 years to 35+ years. The majority of participants 128 (33,6%), were in the 20_24 age group. Of the respondents 291 (76.4%) were married, 112 (29,4%) committed to protestant religion, 238 (62,5%) had no educational level, 305 (80,1%) were unemployed, 288 (75,6%) had a monthly household income below 3500 Mt, which is under half of minimum national salary. 345 (90,6%), had planned their pregnancy, 174 (45,7%) had 3 or more children, 283 (74,3%) depends on their partners or other inmates for decision on family planning. Most of participants 307 (80,6%), have no means of transport to go to health facility and tend to walk, and 120 (31,5%), last about an hour to reach the nearest health facility. Of the participants partners, 193 (53,3%) had no educational level and 225 (62,2%), were unemployed.

### Knowledge on maternal health

Knowledge of participants was assessed by asking participants a range of basic topics on maternal health. These topics are commonly shared during their visits on maternal services, specifically during ANC visits and PNC visits, on the basis of health education and covers but not limited to following: *maternal health; The importance of ANC; the number of visits and time to seek health care after pregnancy; partner participation on maternal health, birth signs, birth preparedness, dangerous signs during perinatal period, PNC and the importance, source of health information and satisfaction with services and care provided during last pregnancy*.

Any response on the correct option was considered good knowledge, any response on the incorrect option was considered weak knowledge.

In general, more than half were marked as correct responses. These statements were then categorized to be included in the regression model. The non statistically significant factors were then used to help explain some of the significant and trend predictors. Table 2 below presents results on participants knowledge and satisfaction with maternal health services

**Table. 2.** Participants knowledge about maternal health and satisfaction with maternal services.

| **Variables categories** | **Frequency** | **percentage** |
| --- | --- | --- |
| **Have you ever heard about maternal health (n=381)** | **n** | **%** |
| Good knowledge | 333 | 87,4% |
| Less knowledge | 48 | 12,6% |
| **Knowledge about the frequency of ANC visits (n=381)** |  |  |
| Good knowledge | 254 | 66,7% |
| Less knowledge | 127 | 33,3% |
| **Opinion about partner attendance on ANC (n=381)** |  |  |
| Good knowledge | 330 | 86,6% |
| Less knowledge | 51 | 13,4% |
| **knowledge about vaginal discharge (bleeding) during pregnancy? (n=381)** |  |  |
| Good knowledge | 348 | 91,3% |
| Less knowledge | 33 | 8,7% |
| **knowledge about vaginal discharge (n=381)** |  |  |
| Good knowledge | 348 | 91,3% |
| Less knowledge | 33 | 8,7% |
| **knowledge about dizziness during pregnancy (n=381)** |  |  |
| Good knowledge | 340 | 89,2% |
| Less knowledge | 41 | 10,8% |
| **knowledge about headaches during pregnancy (n=381)** |  |  |
| Good knowledge | 341 | 89,5% |
| Less knowledge | 40 | 10,5% |
| **knowledge about chest pain during pregnancy (n=381)** |  |  |
| Good knowledge | 345 | 90,6% |
| Less knowledge | 36 | 9,4% |
| **knowledge about belly pain during pregnancy (n=381)** |  |  |
| Good knowledge | 352 | 92,4% |
| Less knowledge | 29 | 7,6% |
| **Satisfaction with ANC (n=375)** |  |  |
| Good knowledge | 318 | 84,8% |
| Less knowledge | 57 | 15,2% |
| **Satisfaction with delivery (n=333)** |  |  |
| Good knowledge | 270 | 81,1% |
| Less knowledge | 63 | 18,9% |
| **Satisfaction with PNC (n=381)** |  |  |
| Good knowledge | 320 | 84,0% |
| Less knowledge | 61 | 16,0% |

### Prevalence of low utilization of maternal services

For our best understanding, we categorized all those above the average as low utilization. For example, ANC visits less than four were categorized as low utilization of ANC services. All those first ANC visits after the first trimester were categorized as low utilization of ANC visits in the first trimester. All those not using institutional delivery were categorized as low utilization of institutional delivery. All those with less than three PNC visits following the 42 days after birth were categorized as low utilization of PNC services.

#### Number of ANC visits

Of the 381 participants 283 (74,3%), (95% CI: 69.7–78.5) had four or more ANC visits, and 98 (25,7%) had less than four ANC visits, during their last pregnancy.

#### Time of first ANC visit

More than a half of participants 211 (56,4%) initiated ANC visit after the first trimester of pregnancy, and 163 (43,6%) (95% CI: 38.6–48.6) within the first trimester.

#### Utilization of institutional delivery

Of the 381 participants, who had their last birth in the year before data collection 267 (70.1%) (95% CI: 65.4–74.5) had delivered at health facility and 117(29.9%) delivered outside health facility

#### Number of PNC visits following 42 days after birth

Of the 381 respondents, 275 (72,2%) (95% CI: 65.4–74.5) had three or more PNC, and 106 (27,8%), had less than three PNC visits. Table 3. Presents the prevalence of low utilization of maternal services.

**Table 3.** Prevalence of low utilization of maternal services.

| Variables categories | Frequency | percentage | IC 95% |
| --- | --- | --- | --- |
| **Number of ANC visit (n=381)** |  |  |  |
| Four or more ANC visit | 283 | 74.3% | 69.7% _ 78.5% |
| Less than four ANC visit | 98 | 25.7% | 21.5% _ 30.3% |
| **Time of first ANC visit (n=374)** |  |  |  |
| Within first trimester | 163 | 43.6% | 38.6% _ 48.6% |
| after first trimester | 211 | 56.4% | 51.4% _ 61,4% |
| **Institutional delivery (n=381)** |  |  |  |
| had delivered at health facility | 267 | 70.1% | 65.4% _ 74.5% |
| delivered outside health facility | 114 | 29.9% | 25.5% _ 34.6% |
| **Number of PNC visits within 42 days after delivery (n=381)** |  |  |  |
| had three or more PNC visits | 275 | 72.2% | 67.5% - 76.5% |
| Had less than three PNC visits | 106 | 27.8% | 23.5% - 32.5% |

#### Reasons for low utilization of maternal services

S2_S5 presents graphics with the reported reasons for not attending the services. The reasons were accessed by asking to follow up questions for those who reported less than the use of four or more ANC visits, first ANC visit after the first trimester, non-institutional delivery, and less than three PNC visits (S2_S5 Figs), respectively.

### Factors associated with low utilization of maternal services during pregnancy, delivery and post delivery

To explore which factors were associated with low utilization of maternal services, we built a simple logistic regression model analysis considering utilization of maternal services as the outcome variable (0=no and 1=yes) and the age, religion, education, employment status, household income, time to reach health centre, means to go to health centre, number of children, partner educational and employment status, knowledge, and satisfaction as predictors variables. Those not significant in the simple model analysis were excluded and the significant variables were then put in the multivariable analysis. Tables 4, 5, 6 and 7 present the results on regression analysis of demographic, knowledge, satisfaction and the use of less than four ANC visits, first ANC visits after the first trimester, non-institutional delivery and less than three PNC visits, respectively.

**Table 4.**
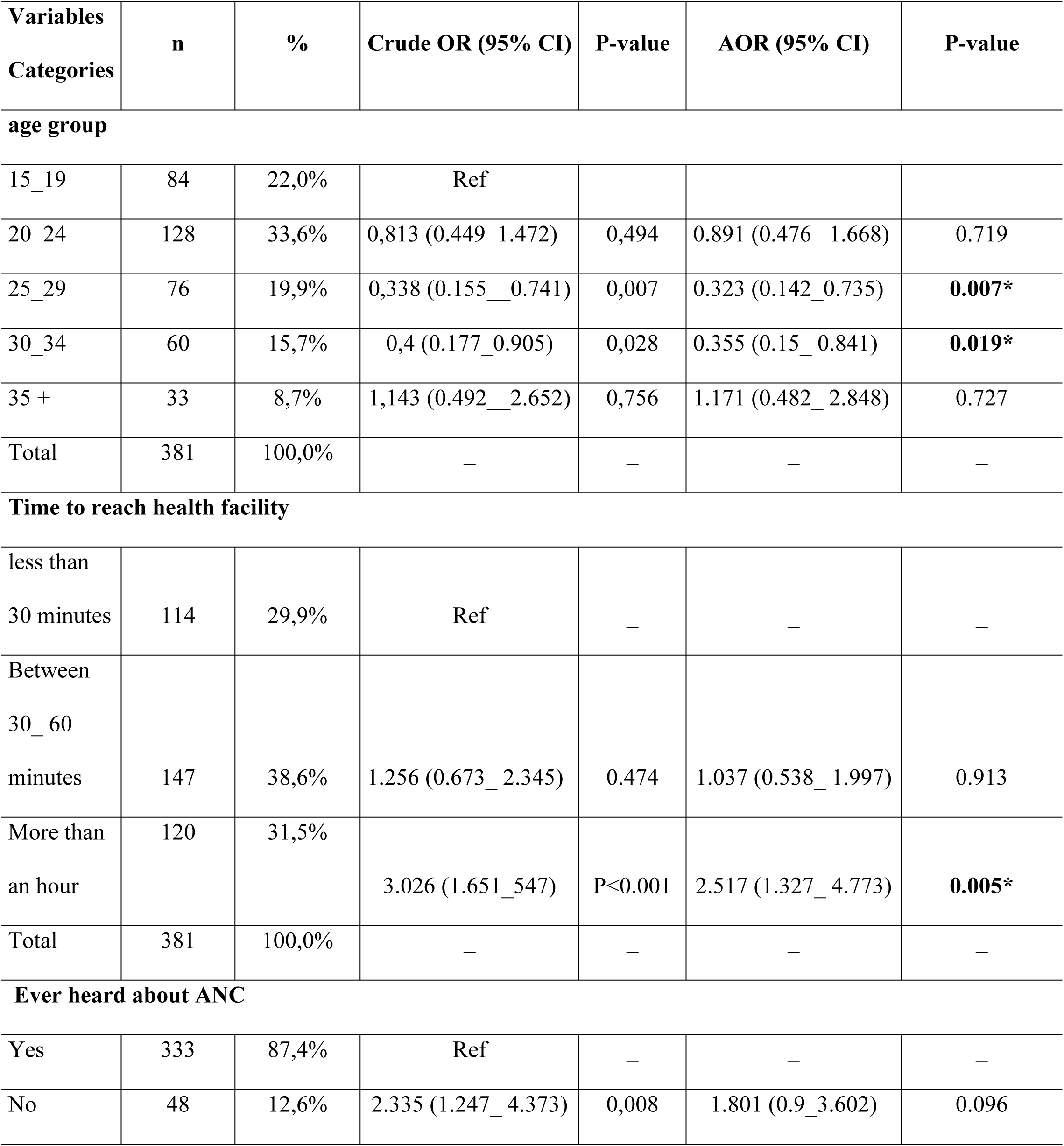
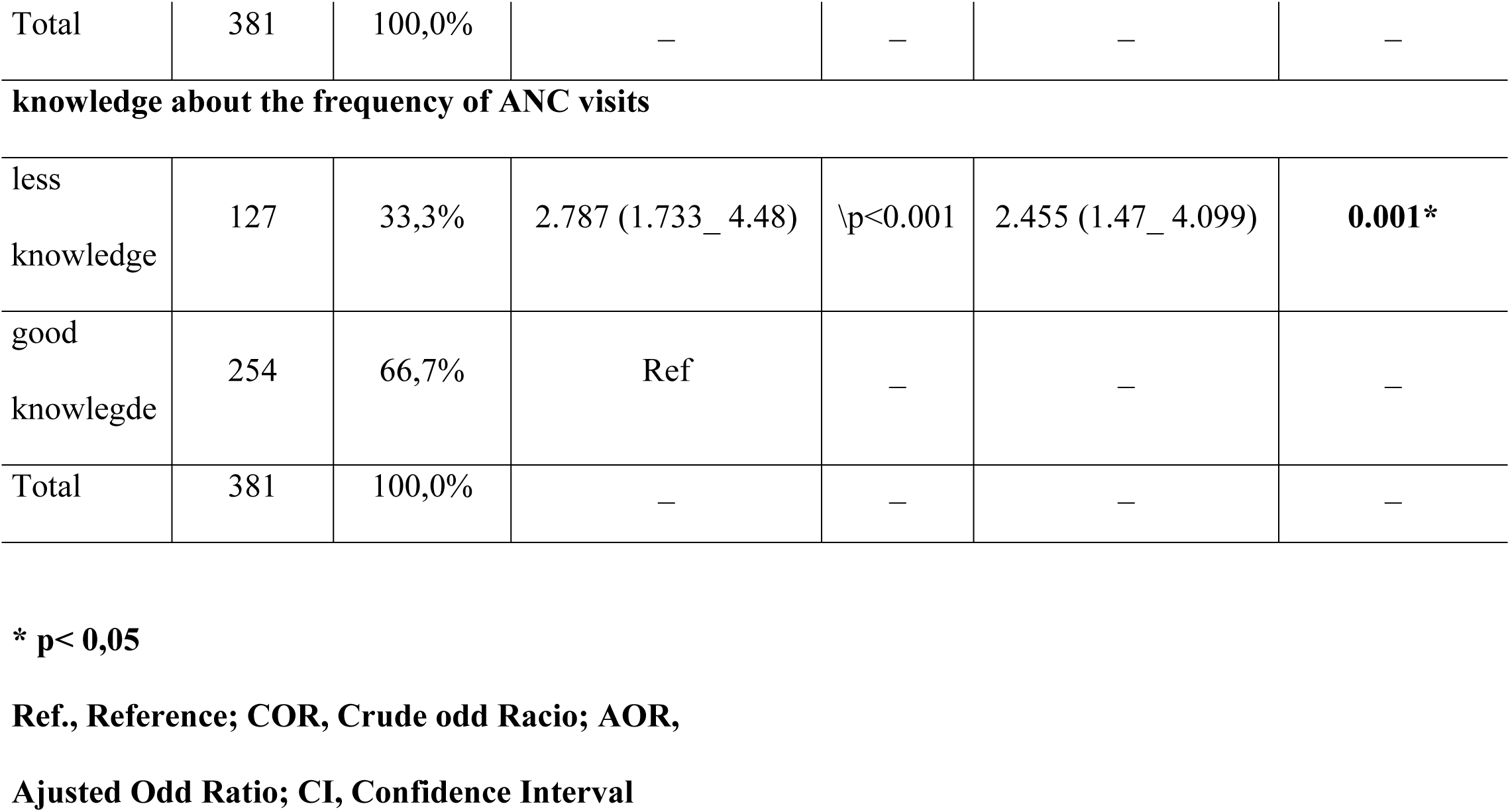
Logistic regression analysis of demographic, knowledge, satisfaction and the use of less than four ANC visits.

| **Variables**<br>**Categories** | **n** | **%** | **Crude OR (95% CI)** | **P-value** | **AOR (95% CI)** | **P-value** |
| --- | --- | --- | --- | --- | --- | --- |
| **age group** |  |  |  |  |  |  |
| 15_19 | 84 | 22,0% | Ref |  |  |  |
| 20_24 | 128 | 33,6% | 0,813 (0.449_1.472) | 0,494 | 0.891 (0.476_ 1.668) | 0.719 |
| 25_29 | 76 | 19,9% | 0,338 (0.155__0.741) | 0,007 | 0.323 (0.142_0.735) | **0.007\*** |
| 30_34 | 60 | 15,7% | 0,4 (0.177_0.905) | 0,028 | 0.355 (0.15_ 0.841) | **0.019\*** |
| 35 + | 33 | 8,7% | 1,143 (0.492__2.652) | 0,756 | 1.171 (0.482_ 2.848) | 0.727 |
| Total | 381 | 100,0% | — | — | — | — |
| **Time to reach health facility** |  |  |  |  |  |  |
| less than<br>30 minutes | 114 | 29,9% | Ref | — | — | — |
| Between<br>30_ 60<br>minutes | 147 | 38,6% | 1.256 (0.673_ 2.345) | 0.474 | 1.037 (0.538_ 1.997) | 0.913 |
| More than<br>an hour | 120 | 31,5% | 3.026 (1.651_547) | P<0.001 | 2.517 (1.327_ 4.773) | **0.005\*** |
| Total | 381 | 100,0% | — | — | — | — |
| **Ever heard about ANC** |  |  |  |  |  |  |
| Yes | 333 | 87,4% | Ref | — | — | — |
| No | 48 | 12,6% | 2.335 (1.247_ 4.373) | 0,008 | 1.801 (0.9_3.602) | 0.096 |
| Total | 381 | 100,0% | — | — | — | — |
| **knowledge about the frequency of ANC visits** |  |  |  |  |  |  |
| less knowledge | 127 | 33,3% | 2.787 (1.733_ 4.48) | \p<0.001 | 2.455 (1.47_ 4.099) | **0.001\*** |
| good knowlegde | 254 | 66,7% | Ref | — | — | — |
| Total | 381 | 100,0% | — | — | — | — |

\* p< 0,05
Ref., Reference; COR, Crude odd Racio; AOR,
Ajusted Odd Ratio; CI, Confidence Interval

**Table 5.** Logistic regression analysis of demographic, knowledge, satisfaction and the use of first ANC visit after the first trimester.

| Variables |  |  |  |  |  |  |
| --- | --- | --- | --- | --- | --- | --- |
| Categories | n | % | Crude OR (95%) | P-value | AOR (95% CI) | Pvalue |
| **Age groups** |  |  |  |  |  |  |
| 15_19 | 82 | 21,9% | Ref | – | – | – |
| 20_24 | 124 | 33,2% | 0.499 (0.277_ 0.898) | 0.020 | 0.509 (0.282_0.919) | **0.025\*** |
| 25_29 | 76 | 20,3% | 0.319 (0.166_ 0.614) | 0.001 | 0,322 (0.167_0.622) | **0.001\*** |
| 30_34 | 59 | 15,8% | 0.52 (0.259_1.042) | 0.065 | 0,522 (0.26_ 1.051) | 0.069 |
| 35 + | 33 | 8,8% | 1.17 (0.476_ 2.873) | 0.733 | 1,204 (0.488_2.972) | 0.687 |
| Total | 374 | 100,0% | – | – | – | – |
| **How often should a pregnant woman go to the ABC during pregnancy** |  |  |  |  |  |  |
| less knowledge | 122 | 32,6% | 1.586 (1.017_ 2.474) | 0.042* | 1.581 (1.003_2.491) | **0.048\*** |
| good knowlegde | 252 | 67,4% | Ref | – | – | – |
| Total | 374 | 100,0% | – | – | – | – |

\* p< 0,05; Ref.,Reference; COR, Crude odd Racio; AOR, Ajusted Odd Ratio; CI, Confidence Interval

#### Logistic regression analysis of demographic, knowledge, satisfaction and the use of less than four ANC visits

In the simple model, the odds of not attending four ANC visits were lower among women with ages between 25_29 (COR, 0. 338, 95% CI: 0.155−0.741, p = 0.002), and the ages between 30_34 (COR, 0.4, 95% CI: 0.177−0.905, p = 0.028) compared to those in the younger and older age categories. The odds ratio of attending less than four ANC were significantly higher among participants who take more than an hour to reach the nearest health facility (COR, 3. 026, 95% CI: 1.651−1.547, p = 0.001) and those with less knowledge about ANC (COR, 2. 335, 95% CI: 1.247−4.373, p = 0.008) and those with less knowledge about the number of ANC visits (OR, 2. 787, 95% CI: 1.733−4.48, p = 0.001). Our multivariable analysis confirmed association between the less than four ANC visits and the middle age of women of 25_29 and 30_34 years old, respectively (AOR, 0. 007, 95% CI: 0.142−0.735, p = 0.007), and (AOR, 0. 355, 95% CI: 0.15−0.841, p = 0.019). Women who delay more than hour to reach the nearest health facility (AOR, 2. 517, 95% CI: 1.327−4.773, p = 0.005), and those with weak knowledge on the frequency of ANC visits (AOR, 2. 455, 95% CI: 1.47−4.099, p = 0.001) are two times more likely to not have the minimum four ANC visits compared to their counterparts who delay less and who have strong knowledge, about the frequency of ANC visits, respectively. There were strong trends that the risk of not attending four ANC visits increased if women was less knowledge about ANC (AOR, 1. 801, 95% CI: 0.9−3.602, p = 0.096) (table 5).

#### Logistic regression analysis of demographic, knowledge, satisfaction and the use of first ANC visit after the first trimester

In the simple model the odds of not attending first ANC visit in the first trimester were lower among women with ages between 25_24 (COR, 0. 499, 95% CI: 0.277−0.898, p = 0.020), those with ages between 25_29 (COR, 0.319, 95% CI: 0.166−0.614, p = 0.001) as well as those with the ages between 20_34 (COR, 0.52, 95% CI: 0.259−1.042, p = 0.065) compared to those in the younger and older age categories. The odds ratio of not attending first ANC visit in the first trimester was significantly higher among participants with less knowledge about the number of ANC visits (COR, 1. 586, 95% CI: 1.017−2.474, p = 0.042). The multivariable analysis confirmed association between not attending first ANC visits in the first trimester and the middle age of mothers (AOR, 0. 509, 95% CI: 0.282−0.919, p = 0.025), and (AOR, 0. 322, 95% CI: 0.167−0.622, p = 0.001) and with weak knowledge about the frequency of ANC visits (AOR, 1. 581, 95% CI: 1.003−2.491, p = 0.048). (Table 6.)

**Table 6.**
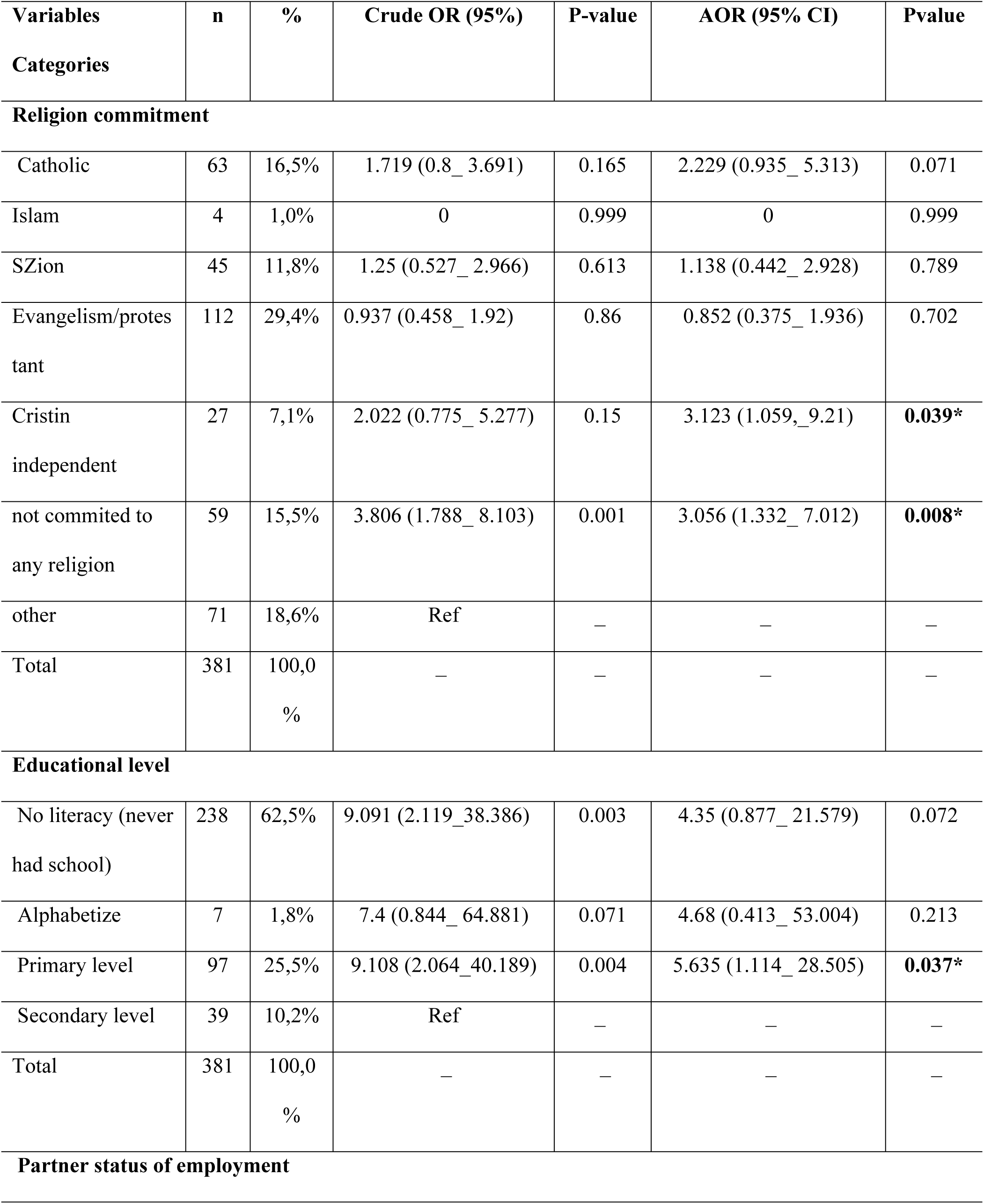
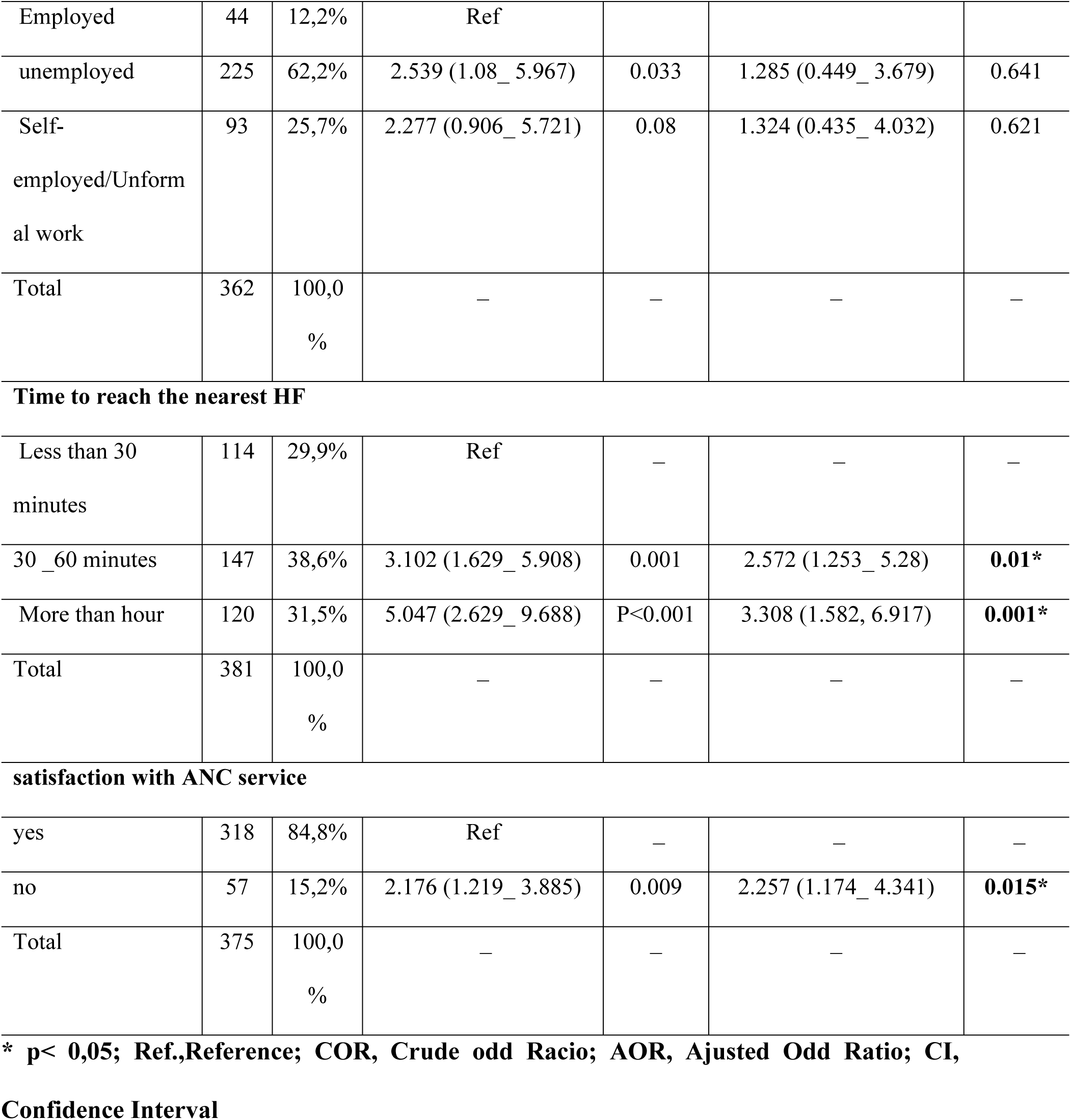
Logistic regression analysis of demographic, knowledge, satisfaction and non-use of institutional delivery.

| Variables<br>Categories | n | % | Crude OR (95%) | P-value | AOR (95% CI) | Pvalue |
| --- | --- | --- | --- | --- | --- | --- |
| **Religion commitment** |  |  |  |  |  |  |
| Catholic | 63 | 16,5% | 1.719 (0.8_ 3.691) | 0.165 | 2.229 (0.935_ 5.313) | 0.071 |
| Islam | 4 | 1,0% | 0 | 0.999 | 0 | 0.999 |
| SZion | 45 | 11,8% | 1.25 (0.527_ 2.966) | 0.613 | 1.138 (0.442_ 2.928) | 0.789 |
| Evangelism/protes<br>tant | 112 | 29,4% | 0.937 (0.458_ 1.92) | 0.86 | 0.852 (0.375_ 1.936) | 0.702 |
| Cristin<br>independent | 27 | 7,1% | 2.022 (0.775_ 5.277) | 0.15 | 3.123 (1.059,_9.21) | **0.039\*** |
| not committed to<br>any religion | 59 | 15,5% | 3.806 (1.788_ 8.103) | 0.001 | 3.056 (1.332_ 7.012) | **0.008\*** |
| other | 71 | 18,6% | Ref | — | — | — |
| Total | 381 | 100,0<br>% | — | — | — | — |
| **Educational level** |  |  |  |  |  |  |
| No literacy (never<br>had school) | 238 | 62,5% | 9.091 (2.119_38.386) | 0.003 | 4.35 (0.877_ 21.579) | 0.072 |
| Alphabetize | 7 | 1,8% | 7.4 (0.844_ 64.881) | 0.071 | 4.68 (0.413_ 53.004) | 0.213 |
| Primary level | 97 | 25,5% | 9.108 (2.064_40.189) | 0.004 | 5.635 (1.114_ 28.505) | **0.037\*** |
| Secondary level | 39 | 10,2% | Ref | — | — | — |
| Total | 381 | 100,0<br>% | — | — | — | — |
| **Partner status of employment** |  |  |  |  |  |  |
| Employed | 44 | 12,2% | Ref |  |  |  |
| unemployed | 225 | 62,2% | 2.539 (1.08_ 5.967) | 0.033 | 1.285 (0.449_ 3.679) | 0.641 |
| Self-employed/Unformal work | 93 | 25,7% | 2.277 (0.906_ 5.721) | 0.08 | 1.324 (0.435_ 4.032) | 0.621 |
| Total | 362 | 100,0% | – | – | – | – |
| Time to reach the nearest HF |  |  |  |  |  |  |
| Less than 30 minutes | 114 | 29,9% | Ref | – | – | – |
| 30 _60 minutes | 147 | 38,6% | 3.102 (1.629_ 5.908) | 0.001 | 2.572 (1.253_ 5.28) | **0.01\*** |
| More than hour | 120 | 31,5% | 5.047 (2.629_ 9.688) | P<0.001 | 3.308 (1.582, 6.917) | **0.001\*** |
| Total | 381 | 100,0% | – | – | – | – |
| satisfaction with ANC service |  |  |  |  |  |  |
| yes | 318 | 84,8% | Ref | – | – | – |
| no | 57 | 15,2% | 2.176 (1.219_ 3.885) | 0.009 | 2.257 (1.174_ 4.341) | **0.015\*** |
| Total | 375 | 100,0% | – | – | – | – |

\* p< 0,05; Ref.,Reference; COR, Crude odd Racio; AOR, Ajusted Odd Ratio; CI,
Confidence Interval

#### Logistic regression analysis of demographic, knowledge, satisfaction and not using institutional delivery

In the simple model the odds of not attending institutional delivery were higher among women not committed with any religion (COR, 3. 806, 95% CI: 1.788−8.103, p = 0.001), those with no education (COR, 9. 091, 95% CI: 2.119−38.386, p = 0.003) and primary education (COR, 9. 108, 95% CI: 2.064−40.189, p = 0.004), those whose partners are not employed (COR, 2. 539, 95% CI: 1.08−5.967, p = 0.033), those delaying more than an hour to reach the nearest health facility (COR, 5. 047, 95% CI: 2.629−9.688, p = 0.001) and those not satisfied with ANC services (COR, 2. 176, 95% CI: 1.219−3.885, p = 0.009) compared to their counterparts in the variables categories. The multivariable analysis confirmed association between not attending institutional delivery and the women’s lack of commitment with religion (AOR, 3. 056, 95% CI: 1.332−7.012, p = 0.008), women’s primary education level status (AOR, 5. 635, 95% CI: 1.114−28.505, p = 0.037), women delaying more than an hour to reach the nearest health facility (AOR, 3. 308, 95% CI: 1.582−6.917, p = 0.001) and women not satisfied with ANC services (AOR, 2. 257, 95% CI: 1.174−4.341, p = 0.015). There were trends that the risk of not attending institutional delivery increased if the women’s partner was unemployed (AOR, 1. 285, 95% CI: 0.449−3.679, p = 0.641) (Table 7).

**Table 7.**
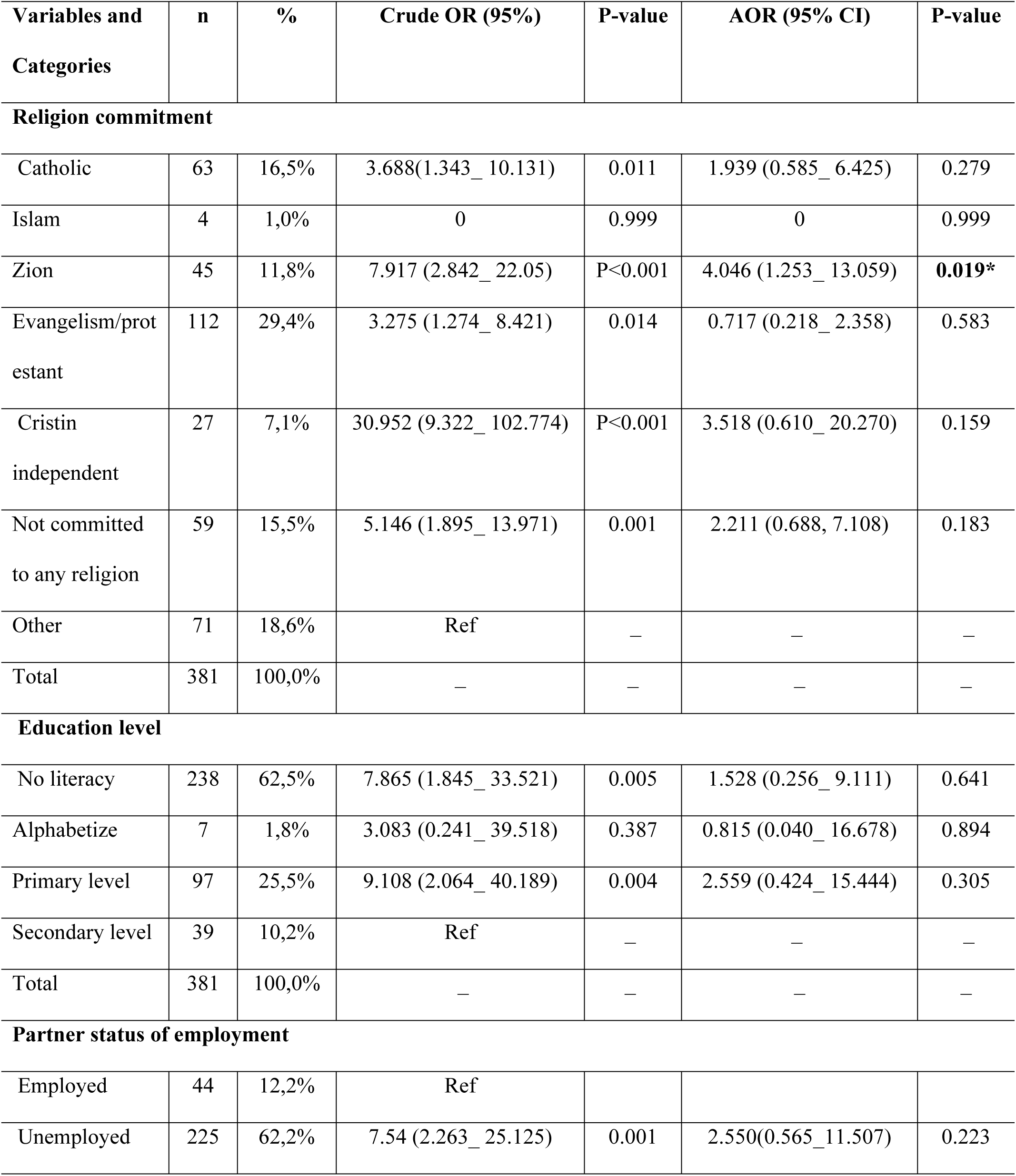
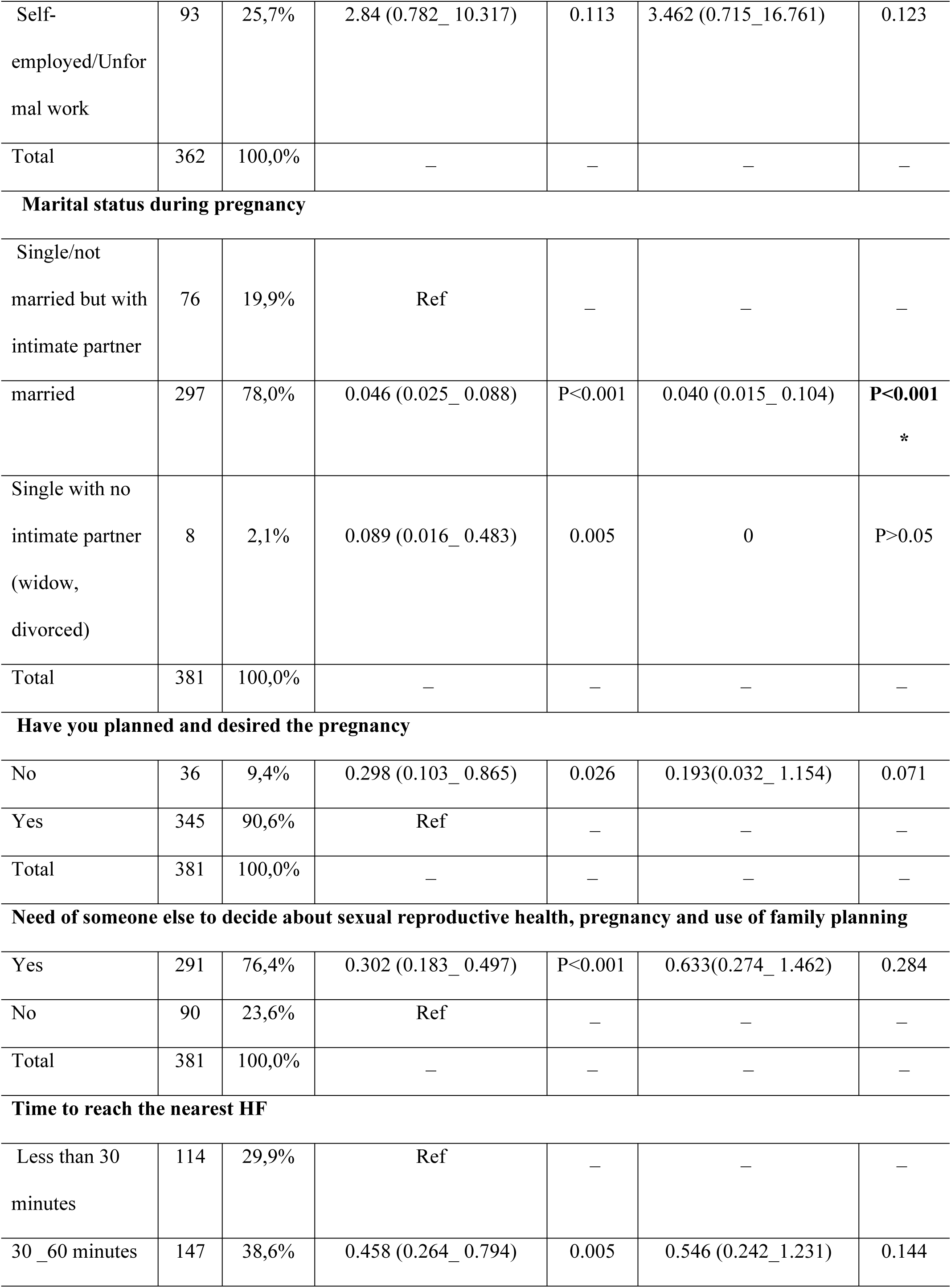
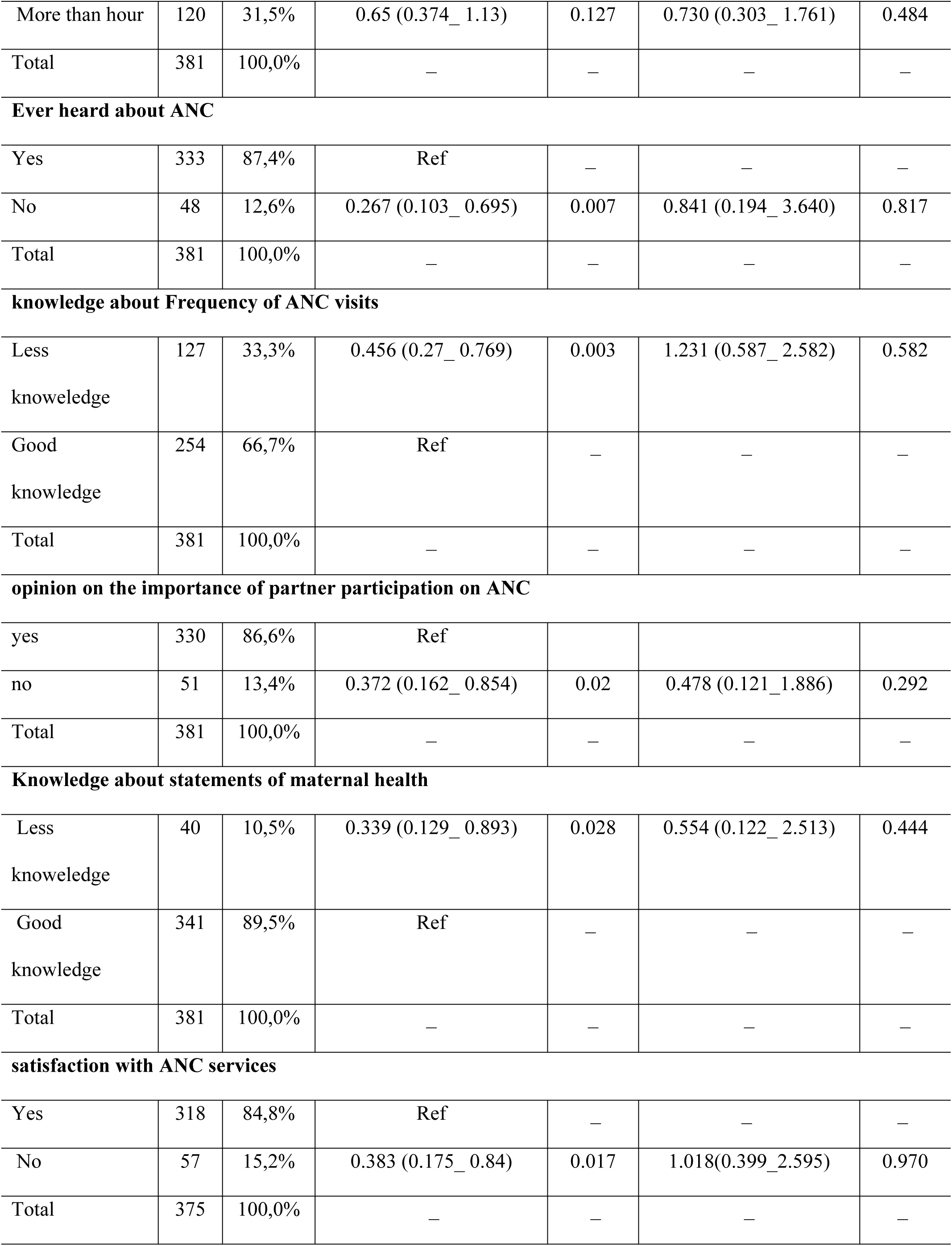
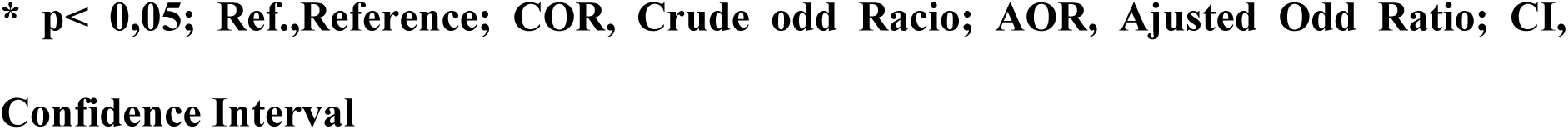
Logistic regression analysis of demographic, knowledge, satisfaction and less than three PNC.

| Variables and Categories | n | % | Crude OR (95%) | P-value | AOR (95% CI) | P-value |
| --- | --- | --- | --- | --- | --- | --- |
| **Religion commitment** |  |  |  |  |  |  |
| Catholic | 63 | 16,5% | 3.688(1.343_ 10.131) | 0.011 | 1.939 (0.585_ 6.425) | 0.279 |
| Islam | 4 | 1,0% | 0 | 0.999 | 0 | 0.999 |
| Zion | 45 | 11,8% | 7.917 (2.842_ 22.05) | P<0.001 | 4.046 (1.253_ 13.059) | **0.019\*** |
| Evangelism/protestant | 112 | 29,4% | 3.275 (1.274_ 8.421) | 0.014 | 0.717 (0.218_ 2.358) | 0.583 |
| Cristin independent | 27 | 7,1% | 30.952 (9.322_ 102.774) | P<0.001 | 3.518 (0.610_ 20.270) | 0.159 |
| Not committed to any religion | 59 | 15,5% | 5.146 (1.895_ 13.971) | 0.001 | 2.211 (0.688, 7.108) | 0.183 |
| Other | 71 | 18,6% | Ref | — | — | — |
| Total | 381 | 100,0% | — | — | — | — |
| **Education level** |  |  |  |  |  |  |
| No literacy | 238 | 62,5% | 7.865 (1.845_ 33.521) | 0.005 | 1.528 (0.256_ 9.111) | 0.641 |
| Alphabetize | 7 | 1,8% | 3.083 (0.241_ 39.518) | 0.387 | 0.815 (0.040_ 16.678) | 0.894 |
| Primary level | 97 | 25,5% | 9.108 (2.064_ 40.189) | 0.004 | 2.559 (0.424_ 15.444) | 0.305 |
| Secondary level | 39 | 10,2% | Ref | — | — | — |
| Total | 381 | 100,0% | — | — | — | — |
| **Partner status of employment** |  |  |  |  |  |  |
| Employed | 44 | 12,2% | Ref |  |  |  |
| Unemployed | 225 | 62,2% | 7.54 (2.263_ 25.125) | 0.001 | 2.550(0.565_11.507) | 0.223 |
| Self-employed/Unfor<br>mal work | 93 | 25,7% | 2.84 (0.782_ 10.317) | 0.113 | 3.462 (0.715_16.761) | 0.123 |
| Total | 362 | 100,0% | — | — | — | — |
| **Marital status during pregnancy** |  |  |  |  |  |  |
| Single/not<br>married but with<br>intimate partner | 76 | 19,9% | Ref | — | — | — |
| married | 297 | 78,0% | 0.046 (0.025_ 0.088) | P<0.001 | 0.040 (0.015_ 0.104) | **P<0.001**<br>* |
| Single with no<br>intimate partner<br>(widow,<br>divorced) | 8 | 2,1% | 0.089 (0.016_ 0.483) | 0.005 | 0 | P>0.05 |
| Total | 381 | 100,0% | — | — | — | — |
| **Have you planned and desired the pregnancy** |  |  |  |  |  |  |
| No | 36 | 9,4% | 0.298 (0.103_ 0.865) | 0.026 | 0.193(0.032_ 1.154) | 0.071 |
| Yes | 345 | 90,6% | Ref | — | — | — |
| Total | 381 | 100,0% | — | — | — | — |
| **Need of someone else to decide about sexual reproductive health, pregnancy and use of family planning** |  |  |  |  |  |  |
| Yes | 291 | 76,4% | 0.302 (0.183_ 0.497) | P<0.001 | 0.633(0.274_ 1.462) | 0.284 |
| No | 90 | 23,6% | Ref | — | — | — |
| Total | 381 | 100,0% | — | — | — | — |
| **Time to reach the nearest HF** |  |  |  |  |  |  |
| Less than 30<br>minutes | 114 | 29,9% | Ref | — | — | — |
| 30 _60 minutes | 147 | 38,6% | 0.458 (0.264_ 0.794) | 0.005 | 0.546 (0.242_1.231) | 0.144 |
| More than hour | 120 | 31,5% | 0.65 (0.374_ 1.13) | 0.127 | 0.730 (0.303_ 1.761) | 0.484 |
| Total | 381 | 100,0% | – | – | – | – |
| **Ever heard about ANC** |  |  |  |  |  |  |
| Yes | 333 | 87,4% | Ref | – | – | – |
| No | 48 | 12,6% | 0.267 (0.103_ 0.695) | 0.007 | 0.841 (0.194_ 3.640) | 0.817 |
| Total | 381 | 100,0% | – | – | – | – |
| **knowledge about Frequency of ANC visits** |  |  |  |  |  |  |
| Less knoweledge | 127 | 33,3% | 0.456 (0.27_ 0.769) | 0.003 | 1.231 (0.587_ 2.582) | 0.582 |
| Good knowledge | 254 | 66,7% | Ref | – | – | – |
| Total | 381 | 100,0% | – | – | – | – |
| **opinion on the importance of partner participation on ANC** |  |  |  |  |  |  |
| yes | 330 | 86,6% | Ref |  |  |  |
| no | 51 | 13,4% | 0.372 (0.162_ 0.854) | 0.02 | 0.478 (0.121_ 1.886) | 0.292 |
| Total | 381 | 100,0% | – | – | – | – |
| **Knowledge about statements of maternal health** |  |  |  |  |  |  |
| Less knoweledge | 40 | 10,5% | 0.339 (0.129_ 0.893) | 0.028 | 0.554 (0.122_ 2.513) | 0.444 |
| Good knowledge | 341 | 89,5% | Ref | – | – | – |
| Total | 381 | 100,0% | – | – | – | – |
| **satisfaction with ANC services** |  |  |  |  |  |  |
| Yes | 318 | 84,8% | Ref | – | – | – |
| No | 57 | 15,2% | 0.383 (0.175_ 0.84) | 0.017 | 1.018(0.399_ 2.595) | 0.970 |
| Total | 375 | 100,0% | – | – | – | – |

\* $p < 0,05$ ; Ref.,Reference; COR, Crude odd Racio; AOR, Ajusted Odd Ratio; CI, Confidence Interval

#### Logistic regression analysis of demographic, knowledge, satisfaction and less than three PNC visits

In the simple model, the odds of less than three PNC visits were higher among women committed to Zion religion (COR, 7. 917, 95% CI: 2.842−22.05, p = 0.001), those not committed with any religion (COR, 5. 146, 95% CI: 1.895−13.971, p = 0.001), compared to those with other religions. The odds of less than three PNC visits, were also higher among women with no education (COR, 9. 865, 95% CI: 1.845−33.521, p = 0.005) as well as with women whose partners are not employed (OR, 7. 54, 95% CI: 2.263−25.125, p = 0.001). The odds of less than three PNC visits, were lower among women with a ‘married’ relationship status (COR, 0. 046, 95% CI: 0.025−0.088, p = 0.001), women who reported unintended pregnancy (COR, 0. 298, 95% CI: 0.103−0.65, p = 0.026), women who cannot decide about their sexual and reproductive health (COR, 0. 302, 95% CI: 0.183−0.497, p = 0.001), women who delay nearly an hour to reach the nearest health facility (COR, 0. 454, 95% CI: 0.264−0.794, p = 0.005), those who never heard about maternal health (COR, 0. 267, 95% CI: 0.103−0.695, p = 0.007), those with weak knowledge about the number of ANC visits (COR, 0. 456, 95% CI: 0.27−0.769, p = 0.003), those who don’t believe in partner importance in maternal health (COR, 0. 372, 95% CI: 0.162−0.854, p = 0.02), those with less knowledge about maternal emergencies (COR, 0. 339, 95% CI: 0.129−0.893, p = 0.028) and those not satisfied with ANC services (COR, 0. 383, 95% CI: 0.175−0.84, p = 0.017), compared to their counterparts in the variables categories.

The multivariable analysis confirmed association between less than three PNC visits and the women commitment with Zion religion (AOR, 4. 046, 95% CI: 1.253−13.059, p = 0.019), and women status of relationship married (AOR, 0. 040, 95% CI: 0.015−0.104, p = 0.001) (Table 8).

## Discussion

This study examined the prevalence of low use of the maternal services and the factors associated with it. The results provide new insights about the utilisation of maternal services among women who ever attended maternal services in Tete province. To the best of our knowledge this is the first exploratory research in Mozambique, that has ever-measured factors associated with low use of ANC, ID and PNC services. Previous studies in Mozambique include the National Demographic Survey, which analysed data on the prevalence of four or more ANC visits, ID e PNC visits [4]. The National survey did not explore the risk factors associated with low use of maternal services. A study which examined the utilisation of four or more ANC services, in the province of Nampula did not include ID and PNC services [31]. Comparisons between our study with existing national data and other similar studies particularly from low middle income countries may be difficult due to the different methodologies in the analysis approach between studies.

This study presents prevalence of use of maternal services and the factors associated with less than four ANC visits, factors associated with first ANC visit after the first trimester, factors associated with not using ID and factors associated with less than three PNC visits.

### Prevalence of low utilization of maternal services

Our results show that prevalence of less than four ANC visits was 25.7%, the prevalence of first ANC visit after the first trimester was 56.4%, prevalence of non-ID was 29.1% and prevalence of less than three PNC 27,8%. Furthermore, prevalence of four or more visits was 74,3%; Prevalence of first ANC within three months pregnancy was 43.6%; Prevalence of institutional delivery was 70.1% and prevalence of three or more PNC was 72.2%.

The prevalence of utilization of maternal services in our study was higher than the results from the previous studies undertaken in Mozambique. The national health demographic survey indicated that of the 49% of women who had four or more ANC, 65% had ID and 36% had attended at least one PNC services [4]. Our findings are quite similar to findings from the study undertaken in Nampula province, where the prevalence of less than four ANC was 50,9% and reported that more than half (60,1%) of women had first their ANC visit after the first trimester. [31].

Our results are similar to studies from low-income countries where the use of ANC, ID and PNC varied from 55.2%, 78.8%, and 40%[30], respectively, but different for the example, from Ethiopia, where a high prevalence of 70% for home deliveries was reported [40], and from countries around SSA and Guinea, where only 19% of women attended at least four ANC and 7% attended PNC [41, 42].

Our findings are also similar to a study undertaken in SSA, among young women. The study reported low utilization of ANC, ID and PNC at 30.1%, 32.3%, and 8.4%, respectively [30]. Similar results were reported in studies from Nigeria, where the use of ANC, ID and PNC was 60,3%, 43,4% and 41,2 %, respectively [29, 43], and similar to those from Sierra Leone for ANC reported at (90.3%), in Congo the ID was reported at (97.5%), and in Cameron, the use of PNC was (88.4%), [44–47].

In general, we observed variations in the prevalences of specific maternal services within countries. The differences in the findings between our study and other may be due differences in the methods used. Most of studies used national representative data from demographic surveys. The methods used and considerations in utilization of maternal services may have also been different. For example, we classified low utilization of ANC as those who attended less than four ANC visits, including those who never utilized these services. Others classified low utilisation of ANC services only to those who never used or those who utilized these only once. Non-ID was deemed as every delivery outside a health facility, while others have classified specific community structures with health providers as ID. Most of studies we use for comparison only examined the use of PNC services one or two times within the 24 or 48 hours after the delivery. In our study we considered the use of PNC services during the 42 days following the delivery and the number of ever visits since.

In the specific context where our study was undertaken, we found high prevalence of utilization of ANC, ID and PNC. Although this may be attributed to local context and use of neighbour maternal services by communities around bordering countries which are much closer. However, the prevalence of low utilisation of four or more ANC in the setting is of much concern, Since in rural settings such as where the study was undertaken, challenges concerning weak socio cultural and health systems structures still prevail and may underpin important determinants of maternal health [33, 48]. The reported reasons for not using ANC, ID and PNC, was attributed to anticipating the physical changes associated with pregnancy, having no reason to do so many visits, not having company, not having money for transport, not satisfied with service not realizing the pregnancy, long distance between residences and health facility, lack of knowledge about the signs of birth, social and cultural construction about the care of newborns. Although we did not include these variables in our regression model, these results should have been interpreted in relation to specific rural context where the study was undertaken and to add and help explain the trends and the statistically significant results reported in our multivariable logistic regression.

The low utilization of maternal services reported in this study may however be the result of a complex cultural environmental and the economic context. A qualitative study conducted in south region in Mozambique, to explore the relationship between health determinants and maternal health, a diverse of socio-cultural, environmental, economic and political factors, showed to be driving the community perceptions on the determinants of maternal health [27, 33]. The reports on less than four ANC attendance, the women initiating ANC visits after the first trimester of pregnancy, the birth deliveries outside health facilities and the less than three PNC attendance reported in this study are of major concern. It seems that the lack of evidence in the reduction on the actual maternal deaths rates in Mozambique is linked to poor quality maternal health care during the perinatal period [18, 27, 31, 35]. Notwithstanding, regular attendance of ANC appointments is proven to be a strategy to accelerate the reductions in maternal mortality [21]. ANC offer opportunities to promote health, care and treatment and to prevent in the early stages, the common dangerous conditions that cause maternal deaths [11, 20, 36]. Women attending more ANC visits are more likely to use institutional delivery and to attend PNC [41].

### Factors associated with low utilization of maternal services

The results of our multivariable analyses showed that women in the middle of reproductive ages of 25 -34 years were less likely to use four or more ANC and to visit ANC in the first trimester than their younger and older counterparts. Delaying more than one hour to reach the nearest health facility, increased the risk to less ANC compared to those delaying less than hour. Weak knowledge about maternal health increased the risk of less ANC visits and with first ANC visit in the first trimester than those with strong knowledge. Women not committed to any religion increased the risk of not utilizing ID. Holding primary education level increased the risk of not utilizing ID than those holding secondary education. Mothers who delay more than an hour and those not satisfied with maternal services in their last experience were more likely to not utilizing ID than those delaying less and those satisfied. Being committed to the Zion religion increased the odds of less PNC visits than those committed to other religions or not committed to any religion. Being married reduced the odds of less PNC visits than the counterparts who were single at the time of data collection. Lastly, there were trends that women whose partners are unemployed are more likely to not use institutional delivery.

There are limited studies conducted in Mozambique to explore factors associated with utilisation of maternal services. Our results on the factors associated with low utilisation of maternal services are similar to most studies done in different regions of Mozambique. In a study conducted in the province of Nampula, findings revealed association between high education, employment and having partner with an increased use of ANC services, including attendance of first visit within the first trimester of pregnancy [31]. A study conducted in Gaza province, reported determinates of economic, sociocultural, environmental factors influencing access and use of maternal services [33]. Similar findings were reported in studies undertaken in Brazil, Nigeria, Ghana, Ethiopia, and elsewhere [8, 11, 28, 29, 40, 43].

The results on the factors associated with low use of Maternal services are similar with most studies conducted around settings in low, middle and high-income countries [6, 49, 50].

There are diverse possibilities to explain relationships between the determinants affecting the utilisation of maternal health services and the results reported in our study given the context.

On one hand, the results on trends of unemployment status and low education in study participants may have reduced the ability of women and their partners to understand the importance and needs for utilizing maternal services [31, 51–54]. On the other hand, the high unemployment rates in the country [55, 56] may be extended to the region and affected the population of the study. In this study, our binary regression showed that being unemployed and having partner who is also unemployed, was associated with low utilisation of maternal services. Unemployment status and the low household income, limit the ability of families to acquire the means for their needs, this includes transportation to health facilities during pregnancy, delivery and after delivery [49, 57–61]. The means for transportation in these settings, should be seen as an important factor due to distance between communities and health facilities [18, 27, 31, 33]. Our study revealed that participants who delay an hour to reach the nearest health facility were at an increased risk of low use of institutional delivery, as compared to their counterparts who delay less. It should be noted that in this setting there are no formal transportation systems or structures that facilitate mobility. People usually do walk long distances to reach health facilities. This said, there is an urgent need for Mozambican health system to include programs and coordinated efforts to reduce the barriers for accessing health facilities [27, 33, 62]. In Mozambique, community providers are more concerned on educational activities for prevention and more referral counselling for pregnant women rather than providing treatment and response to emergency needs of women during perinatal period. Looking at Ghana’s experience on the program aiming at promoting maternal service utilization, results reported increased utilization of 4 or more ANC visits and ID through implementing consistent community health education activities and from having introduced parallel systems of referral transportation. [25, 44, 60, 63].

In this study married women were those less likely to less use of PNC services than those who identified as single. Being single and pregnant my affect women in different ways and perspectives. These results may suggest at same time improved autonomy of married women to decide on important maternal health issues and the use of PNC services. In Mozambique, particularly in rural areas, men still have crucial influence and important decisions about various aspects of sexual and reproductive health, including the use of maternal services. As for other sexual reproductive and maternal factors, partners may be the ones who decide on the use of health services, including going to access maternal services [33, 36]. The divergent social and cultural dynamics of the community and families in the setting where the study was undertaken may have impact in the utilization of maternal services among married women.

Moreover, in the bivariate analyses women who rely on partner to decide on family planning and those who reported not having planed or desired the pregnancy, not attributing importance for partner involvement in maternal health were less likely to less use PNC services. However, the need of partner’s approval for family planning may demonstrate the prevailing cultural male dominant norms and the inability of women to decide about their reproductive health.

The rooted sociocultural barriers still prevailing in the rural setting need to be urgently addressed and to change male dominant norms to a more gender sensitive approach, and women empowerment taking in consideration the important and positive influence of male partners in improving maternal health outcomes [33, 51, 64–66].

On the other hand, being single and pregnant may mean increased vulnerability, leading to increase of emotions and anxiety about the pregnancy and the future, and these may lead to more use of health services for care, since the single woman is the only one who decides. Given the local context, these women are more likely to experience increased emotional, social and financial constraints and less likely to attend maternal services, due to the reduced means of support for pregnancy related emergent needs [49, 55, 64–66]. However, these results should be interpreted with caution in different contexts. Our variables on the reason not attending maternal services, one was not having partner. The Socio-cultural and economic aspects around single pregnant women may have its contributions leading to stigma and reduced access to quality maternal services [31, 33, 36, 67–69].

It is important to take into consideration, the important role of men to promote maternal and family health, and the consideration of equitable services where single women receive equal care and feel safe to use their rights of care and health for both mothers and their babies.

In this study the median age of mothers was significantly associated with reduced risk of less ANC compared with very young or much older woman. The age in which women get pregnant seems to be an important factor affecting the ability to access and use maternal services. This may be due to their middle reproductive age, with high fertility and may have had previous experiences of pregnancy and maternal services. Previous negative experiences may reduce the women’s ability to use maternal services in subsequent pregnancies [49, 70]. With the high fertility in rural areas in Mozambique, it is not unusual that woman in the ages between 25-34 may have had previous experiences of pregnancy and have used maternal services [4]. Negative experiences during maternity, for example, long waiting time, poor attitudes with providers and women’s dissatisfaction may impact in reduction of maternal health utilization in subsequent pregnancies. Contrarily, the youngest women, which may be experiencing a first-time pregnancy, with reduced experience about pregnancy, and the need to use health services for information and care may impact their ability to understand the importance of using maternal services. The older adults who may be more experienced and more concerned about pregnancy might use more of the available maternal services, due to their ability to understand the importance and positive impacts of using these. It is important to take into account that the older age group of pregnant women are also under obstetric risks such as haemorrhage, hypertension and other complications and therefore need more of maternal service visits to prevent and monitor these. Interpretating these results need to take into consideration the contextual and personal perspectives, means and attributions around maternity. Interventions should incorporate young and adult women’s specific needs to increase their use and preferences for maternal health services in this setting [71, 72].

This study has revealed association between women’s dissatisfaction with previous maternal services care and less use of ID. Previous studies in Mozambique, undertaken in both rural and urban areas, have reported evidence on provider’s attitudes of disrespect and abuse during maternal care [9, 27, 36]. Similar findings on dissatisfaction with maternal services were reported in studies from SSA and countries outside SSA [55, 70]. Although we have pointed important factors related to low use of maternal services, our study did not include all indicators, and caution should be observed when interpreting the results. We acknowledge other factors may play crucial role on utilization of maternal service. In previous studies there were concerning factors identified that may have impact in woman access to ANC. This includes the health system function and structure, the attitudes, knowledge and skills of providers towards assistance during the perinatal period, and women’s personal factors. These barriers hindered the implementation of recommended ANC package care in Mozambique and influenced the adequate utilisation of ANC, ID and PNC services [4, 27, 31, 35]

## Conclusion

This study demonstrates prevalence of low utilisation of ANC, ID and PNC services. Factors associated with utilization of less than four ANC visits, non-ID, first ANC visit after first trimester and less than three PNC included being in middle of reproductive age, being married, low educational status, delaying an hour to reach the nearest health facility, not satisfied with maternal services, weak knowledge on maternal health, not committed to any religion and being committed to Zion religion. There is an urgent need for strategies to improve access to health services, by removing socio economic, cultural and structural barriers limiting autonomy and ability for women to attend four or more ANC visits, initiating ANC in the first trimester, attending institutional delivery and attending three or more PNC visits.

### Limitations

Our study had several limitations:

- First, the study makes use of a cross-sectional design which is unable to establish causal relationships between the outcome and explanatory variables.
- The sample of the study was taken from maternal health facilities users’ population, and participants were recruited only if they were health facility users. Our findings may not be generalised for general women in Mozambique, neither for other health facilities users.
- This study used an interview survey, based on women’s self-reports of past events. We did not search for other validation sources. This method is likely to bring social desirability and recall bias. This could have affected validity and reliability. However, biases are less likely from maternal health related events, due to reduced sensitivity of the topic. We also included only women attending the maternal services, who had given their last live birth within a year.

### Strengths

- The strength of our study is that the questionnaire was designed from the available literature and the theoretical model to explain the determinants of maternal health services utilization [39]. It was piloted and adapted to the contextual environment.
- This study used exploratory primary data. This is the first study in Mozambique providing simultaneously the prevalence and factors associated with low utilization of the three main maternal services.
- Our study offers important insights to document evidence-based important factors hindering maternal services users to utilize services during pregnancy, birth and after birth. The findings will inform a program designed to improve maternal health to be implemented in a specific selected health facility.

### Implications

This study has provided important information and contributions regarding evidence on the low utilization of maternal health services. The findings may be utilised to inform policy makers for contextual tailored interventions to improve the opportunities for pregnant women and their partners to increase education and paid work so that families and households’ incomes are made possible to arrange for means to access maternal services during pregnancy, birth and after birth.

There is also a need to improve the male involvement initiatives, so that male partners are aware of their importance and responsibility in maternal health. The latter should include families and communities to support and empower married and single pregnant women. This study demonstrates how individual, community, political and structural factors interact to influence the use of maternal services. The challenge over the next six years is to work towards reducing the maternal mortality national rate to adequate levels by 2030. Efforts to improve maternal health and archive the 2030 targets 3.1, 3.8 and 5 should be made at different multilevel sectors.

## Data Availability

The data underlying this article cannot be shared publicly due to the privacy of the individuals that participated in the study. The data will be shared at reasonable request to the corresponding author

## Acknowledgments

The authors would like to thank the respondents and research assistants for their participation. The authors would like to thank the Provincial Coordinator of the Project in Tete, JLM, for his contributions during the research phase. The authors acknowledge the Government of Flanders, for the provision of resources toward this article.

## Supporting information

S1 File. **This is the S1 file interview questionnaire.**

S2 Fig. **This is the S2 Fig reasons for not attending four or more ANC visits**

S3 Fig. **This is the S3 Fig reasons for not attending first ANC in the first trimester**

S4 Fig. **This is the S4 Fig reasons for not attending institutional delivery**

S5 Fig. **This is the S5 Fig reasons for not attending three or more PNC visits**

## References

1. Organization, W.H., Trends in maternal mortality 2000 to 2020: estimates by WHO, UNICEF, UNFPA, World Bank Group and UNDESA/Population Division. Trends in maternal mortality 2000 to 2020: estimates by WHO, UNICEF, UNFPA, World Bank Group and UNDESA/Population Division. Geneva: World Health Organization; 2023. Licence: CC BY-NC-SA 3.0 IGO., 2023. CIP data are available at http://apps.who.int/iris.

2. Saúde, M.d., Relatório Anual de Auditoria de Mortes Maternas e Neonatais– 2017. 2019.

3. (DPC)-, M.d.S.D.d.P.e.C. and M. Departamento de Informação para a Saúde (DIS) – Maputo, ANUÁRIO ESTATÍSTICO DE SAÚDE 2019. 2020.

4. 2022–23., I.N.d.E.I.e.I.I.D.e.d.S.e.M. and M.e.R. Maputo, Maryland, EUA: INE e ICF, Instituto Nacional de Estatística (INE) e ICF. 2023. Inquérito Demográfico e de Saúde em Moçambique 2022–23. Maputo, Moçambique e Rockville, Maryland, EUA: INE e ICF. 2023.

5. Organization, W.h. Target 3.1 Reduce the global maternal mortality ratio to less than 70 per 100 000 live births. 9–11-2023.

6. Rashid, M. and D. Antai, Socioeconomic position as a determinant of maternal healthcare utilization: a population-based study in Namibia. Journal of research in health sciences, 2014. 14(3): p. 187–192.

7. Health, W.H.O.R., Pregnancy, childbirth, postpartum, and newborn care: a guide for essential practice. 2003: World Health Organization.

8. Haruna, U., G. Dandeebo, and S.Z. Galaa, Improving access and utilization of maternal healthcare services through focused antenatal care in rural Ghana: a qualitative study. Advances in Public Health, 2019. 2019.

9. Singh, R., et al., Utilization of maternal health services and its determinants: a cross-sectional study among women in rural Uttar Pradesh, India. Journal of health, population and nutrition, 2019. 38: p. 1–12.

10. Greenberg, R.S., The impact of prenatal care in different social groups. American Journal of Obstetrics and Gynecology, 1983. 145(7): p. 797–801.

11. Khan, S., S.I. Haider, and R. Bakhsh, Socio-economic and cultural determinants of maternal and neonatal mortality in Pakistan. Global Regional Review, 2020. 1: p. 1–7.

12. Miller, S., et al., Quality of care in institutionalized deliveries: the paradox of the Dominican Republic. International Journal of Gynecology & Obstetrics, 2003. 82(1): p. 89–103.

13. Randive, B., V. Diwan, and A. De Costa, India’s conditional cash transfer programme (the JSY) to promote institutional birth: is there an association between institutional birth proportion and maternal mortality? PloS one, 2013. 8(6): p. e67452.

14. Carroli, G., C. Rooney, and J. Villar, How effective is antenatal care in preventing maternal mortality and serious morbidity? An overview of the evidence. Paediatric and perinatal Epidemiology, 2001. 15: p. 1–42.

15. Carroli, G., C. Rooney, and J. Villar, How effective is antenatal care in preventing maternal mortality and serious morbidity. Sheng zhi yu bi yun= Reproduction and Contraception, 2005. 26(5): p. 300–314.

16. Oyerinde, K., Can antenatal care result in significant maternal mortality reduction in developing countries. J Community Med Health Educ, 2013. 3(2): p. 2–3.

17. Fiscella, K., Does prenatal care improve birth outcomes? A critical review. Obstetrics & Gynecology, 1995. 85(3): p. 468–479.

18. Cane, R.M., et al., Structural readiness of health facilities in Mozambique: how is Mozambique positioned to deliver nutrition-specific interventions to women and children? Journal of Global Health Reports, 2023. 7: p. e2023074.

19. Ministério da Saúde, I.N.d.E., ICF Internacional, Inquérito de indicadores de imunização, malária e HIV/SIDA em Moçambique 2015. 2015, INS, INE e ICF International Rockville, Maryland, EUA.

20. nstituto Nacional de Estatística (INE), I.N.d.S.I.e.J.H.U.J. and M.I. Sistema de Vigilância de Eventos Vitais e Causas de Morte (COMSA), INS e JHU., Instituto Nacional de Estatística (INE), Instituto Nacional de Saúde (INS) e Johns Hopkins University (JHU). Sistema de Vigilância de Eventos Vitais e Causas de Morte (COMSA), Moçambique 2019. INE, INS e JHU. 2023.

21. Organization, W.H., WHO recommendations on antenatal care for a positive pregnancy experience: screening, diagnosis and treatment of tuberculosis disease in pregnant women. Evidence-to-action brief: Highlights and key messages from the World Health Organization’s 2016 global recommendations. 2023: World Health Organization.

22. Chavane, L., et al., Implementation of evidence-based antenatal care in Mozambique: a cluster randomized controlled trial: study protocol. BMC health services research, 2014. 14: p. 1–11.

23. https://pt.wikipedia.org/wiki/Tete_(prov%C3%ADncia), https://pt.wikipedia.org/wiki/Tete_(prov%C3%ADncia).2014,https://pt.wikipedia.org/wiki/Tete_(prov%C3%ADncia).

24. Villar, J., et al., WHO antenatal care randomised trial for the evaluation of a new model of routine antenatal care. The lancet, 2001. 357(9268): p. 1551–1564.

25. Lee, H., et al., The impact of the interventions for 4+ antenatal care service utilization in the democratic republic of congo: a decision tree analysis. Annals of Global Health, 2019. 85(1).

26. Villar, J. and P. Bergsjo, WHO antenatal care randomized trial: manual for the implementation of the new model, in WHO Antenatal care randomized trial: manual for the implementation of the new model. 2002. p. 37–37.

27. Biza, A., et al., Challenges and opportunities for implementing evidence-based antenatal care in Mozambique: a qualitative study. BMC Pregnancy and Childbirth, 2015. 15(1): p. 1–10.

28. Okeke, N.C. and I.I. Ofonere, Determinant and utilization of maternal and child health care services among women of reproductive age in Enugu State, Nigeria. International Journal of Health and Social Inquiry, 2023. 9(1).

29. Ademuyiwa, I.Y., et al., Antenatal Care Services Utilization and Factors Influencing it Among Pregnant Women in a Teaching Hospital, Lagos, Nigeria. 2020.

30. Bain, L.E., et al., Prevalence and determinants of maternal healthcare utilisation among young women in sub-Saharan Africa: cross-sectional analyses of demographic and health survey data. BMC public health, 2022. 22(1): p. 647.

31. Reis-Muleva, B., et al., Antenatal care in Mozambique: Number of visits and gestational age at the beginning of antenatal care. Revista latino-americana de enfermagem, 2021. 29.

32. Jaén-Sánchez, N., et al., Adolescent motherhood in Mozambique. Consequences for pregnant women and newborns. Plos one, 2020. 15(6): p. e0233985.

33. Firoz, T., et al., Community perspectives on the determinants of maternal health in rural southern Mozambique: a qualitative study. Reproductive Health, 2016. 13: p. 123–131.

34. Jennings, L., et al., Women’s empowerment and male involvement in antenatal care: analyses of Demographic and Health Surveys (DHS) in selected African countries. BMC pregnancy and childbirth, 2014. 14: p. 1–11.

35. I.C.f.R.H., M.A.K.U.C.o. Excellence, and i.W.a.C. Health, CONTINUITY OF ESSENTIALHEALTH SERVICES STUDY: Exploring the efects of the COVID-19 Pandemic on the demand for and utilisation of maternal, newborn, child health services in Mozambique. 2021.

36. Galle, A., et al., Disrespect and abuse during facility-based childbirth in southern Mozambique: a cross-sectional study. BMC pregnancy and childbirth, 2019. 19: p. 113.

37. https://data.worldbank.org/indicator/SL.UEM.TOTL.FE.ZS, https://data.worldbank.org/indicator/SL.UEM.TOTL.FE.ZS. 20-03-24.

38. Andersen, R. and J.F. Newman, Societal and individual determinants of medical care utilization in the United States. The Milbank Memorial Fund Quarterly. Health and Society, 1973: p. 95–124.

39. Alkhawaldeh, A., et al., Application and Use of Andersen’s Behavioral Model as Theoretical Framework: A Systematic Literature Review from 2012–2021. Iranian Journal of Public Health, 2023. 52(7): p. 1346.

40. Tarekegn, S.M., L.S. Lieberman, and V. Giedraitis, Determinants of maternal health service utilization in Ethiopia: analysis of the 2011 Ethiopian Demographic and Health Survey. BMC pregnancy and childbirth, 2014. 14: p. 1–13.

41. Soubeiga, D., D. Sia, and L. Gauvin, Increasing institutional deliveries among antenatal clients: effect of birth preparedness counselling. Health Policy and Planning, 2014. 29(8): p. 1061–1070.

42. Ahinkorah, B.O., et al., Determinants of antenatal care and skilled birth attendance services utilization among childbearing women in Guinea: evidence from the 2018 Guinea Demographic and Health Survey data. BMC Pregnancy and Childbirth, 2021. 21: p. 1–11.

43. Babalola, S. and A. Fatusi, Determinants of use of maternal health services in Nigeria-looking beyond individual and household factors. BMC pregnancy and childbirth, 2009. 9: p. 1–13.

44. Koroma, M.M., et al., The quality of free antenatal and delivery services in Northern Sierra Leone. Health research policy and systems, 2017. 15: p. 13–20.

45. Vallières, F., et al., Can Sierra Leone maintain the equitable delivery of their Free Health Care Initiative? The case for more contextualised interventions: results of a cross-sectional survey. BMC health services research, 2016. 16: p. 1–12.

46. ML, A.N., et al., Determinants of maternal health services utilization in urban settings of the Democratic Republic of Congo–a case study of Lubumbashi City. BMC pregnancy and childbirth, 2012. 12: p. 1–13.

47. Yaya, S., et al., Predictors of skilled birth attendance among married women in Cameroon: further analysis of 2018 Cameroon Demographic and Health Survey. Reproductive Health, 2021. 18: p. 1–12.

48. Makanga, P.T., et al., Seasonal variation in geographical access to maternal health services in regions of southern Mozambique. International journal of health geographics, 2017. 16(1): p. 1–16.

49. Domingues, R.M.S.M., et al., Access to and utilization of prenatal care services in the Unified Health System of the city of Rio de Janeiro, Brazil. Revista Brasileira de Epidemiologia, 2013. 16: p. 953–965.

50. Kassaw, A., A. Debie, and D.M. Geberu, Quality of prenatal care and associated factors among pregnant women at public health facilities of Wogera District, Northwest Ethiopia. Journal of pregnancy, 2020. 2020.

51. Okedo-Alex, I.N., et al., Determinants of antenatal care utilisation in sub-Saharan Africa: a systematic review. BMJ open, 2019. 9(10): p. e031890.

52. Schultz, T.P., Studying the impact of household economic and community variables on child mortality. population and Development Review, 1984. 10: p. 215–235.

53. Dimbuene, Z.T., et al., Women’s education and utilization of maternal health services in Africa: a multi-country and socioeconomic status analysis. Journal of biosocial science, 2018. 50(6): p. 725–748.

54. Ovikuomagbe, O., Determinants of maternal healthcare utilization in Nigeria. African Research Review, 2017. 11(2): p. 283–294.

55. Namasivayam, A., et al., The role of gender inequities in women’s access to reproductive health care: a population-level study of Namibia, Kenya, Nepal, and India. International journal of women’s health, 2012: p. 351–364.

56. Group, W.B., https://data.worldbank.org/country/mozambique. 2024: https://data.worldbank.org/country/mozambique.

57. Leive, A. and K. Xu, Coping with out-of-pocket health payments: empirical evidence from 15 African countries. Bulletin of the World Health Organization, 2008. 86(11): p. 849–856C.

58. Fagbamigbe, A.F. and E.S. Idemudia, Barriers to antenatal care use in Nigeria: evidences from non-users and implications for maternal health programming. BMC pregnancy and childbirth, 2015. 15: p. 1–10.

59. Arthur, E., Wealth and antenatal care use: implications for maternal health care utilisation in Ghana. Health economics review, 2012. 2: p. 1–8.

60. Amu, H. and D.K. Sekyi, Effects of spatial location and household wealth on the utilisation of skilled birth attendants at delivery among women in Rural Ghana. Ghana Journal of Geography, 2018. 10(1): p. 58–77.

61. Nuamah, G.B., et al., Access and utilization of maternal healthcare in a rural district in the forest belt of Ghana. BMC pregnancy and childbirth, 2019. 19: p. 1–11.

62. Gudu, W. and B. Addo, Factors associated with utilization of skilled service delivery among women in rural Northern Ghana: a cross sectional study. BMC pregnancy and childbirth, 2017. 17: p. 1–10.

63. Byrne, A., et al., What works? Strategies to increase reproductive, maternal and child health in difficult to access mountainous locations: a systematic literature review. PloS one, 2014. 9(2): p. e87683.

64. Kea, A.Z., et al., Exploring barriers to the use of formal maternal health services and priority areas for action in Sidama zone, southern Ethiopia. BMC pregnancy and childbirth, 2018. 18: p. 1–12.

65. Kaba, M., et al., Sociocultural determinants of home delivery in Ethiopia: a qualitative study. International journal of women’s health, 2016: p. 93–102.

66. Kumbani, L., et al., Why some women fail to give birth at health facilities: a qualitative study of women’s perceptions of perinatal care from rural Southern Malawi. Reproductive health, 2013. 10: p. 1–12.

67. Mohammed, S., I. Yakubu, and I. Awal, Sociodemographic factors associated with women’s perspectives on male involvement in antenatal care, labour, and childbirth. Journal of pregnancy, 2020. 2020.

68. Lowe, M., Social and cultural barriers to husbands’ involvement in maternal health in rural Gambia. The Pan African medical journal, 2017. 27.

69. Singh, A., A. Kumar, and P. Pranjali, Utilization of maternal healthcare among adolescent mothers in urban India: evidence from DLHS-3. PeerJ, 2014. 2: p. e592.

70. Reynolds, H.W., E.L. Wong, and H. Tucker, Adolescents’ use of maternal and child health services in developing countries. International family planning perspectives, 2006: p. 6–16.

71. Conde-Agudelo, A., J.M. Belizán, and C. Lammers, Maternal-perinatal morbidity and mortality associated with adolescent pregnancy in Latin America: Cross-sectional study. American journal of obstetrics and gynecology, 2005. 192(2): p. 342–349.

72. Vogel, J.P., et al., Maternal complications and perinatal mortality: findings of the World Health Organization Multicountry Survey on Maternal and Newborn Health. BJOG: An International Journal of Obstetrics & Gynaecology, 2014. 121: p. 76–88.

